# Maternal B-vitamin and vitamin D status before, during and after pregnancy, and the influence of supplementation preconception and during pregnancy: NiPPeR double-blind randomized controlled trial

**DOI:** 10.1101/2023.06.19.23291584

**Authors:** Keith M Godfrey, Philip Titcombe, Sarah El-Heis, Benjamin B Albert, Elizabeth Huiwen Tham, Sheila J Barton, Timothy Kenealy, Mary Foong-Fong Chong, Heidi Nield, Yap Seng Chong, Shiao-Yng Chan, Wayne S Cutfield, NiPPeR Study Group

## Abstract

**Background:** Maternal vitamin status preconception and during pregnancy have important consequences for pregnancy outcome and offspring development. Changes in status from preconception to early and late pregnancy and postpartum have been inferred from cross-sectional data, with lower pregnancy concentrations often ascribed to plasma volume expansion, but without truly longitudinal data from preconception through pregnancy and post-delivery, and sparse data on the influence of supplementation. This study characterized longitudinal patterns of maternal vitamin status from preconception, through early and late pregnancy, to 6-months post-delivery, and determined the influence of supplementation.

**Methods and Findings:** Between 2015-2017, 1729 UK, Singapore and New Zealand women aged 18-38 years planning conception were recruited from the community to a double-blind controlled trial and randomized to a standard (control) or an intervention supplement preconception and throughout pregnancy. Vitamins common to both supplements were folic acid and β-carotene, with the intervention additionally including riboflavin, vitamins B6, B12 and D in amounts available in over-the-counter supplements, alongside iron, calcium and iodine (control and intervention) and myo-inositol, probiotics and zinc (intervention only).

We measured maternal plasma concentrations of B-vitamins, vitamin D and insufficiency/deficiency markers (homocysteine, hydroxykynurenine-ratio, methylmalonic acid), at recruitment and 1-month after commencing intervention preconception, in early and late pregnancy, and post-delivery (6-months after supplement discontinuation). From all timepoint data, we derived standard deviation scores (SDS) to characterize longitudinal changes in controls and differences between control and intervention participants. At recruitment preconception, significant proportions had marginal or low plasma status for folate (29.2% <13.6 nmol/L), riboflavin (7.5% <5 nmol/L, 82.0% ≤26.5 nmol/L), vitamin B12 (9.1% <221 pmol/L) and vitamin D (48.7% <50 nmol/L). Among controls, plasma concentrations showed differing longitudinal patterns from preconception; riboflavin fell through early/late pregnancy, 25-hydroxyvitamin D was unchanged in early pregnancy, and vitamin B6 and B12 concentrations declined through pregnancy, becoming >1 SDS lower than baseline by 28 weeks gestation, with 54.2% developing a low late pregnancy vitamin B6 (pyridoxal 5-phosphate <20 nmol/L). Preconception, the control/intervention groups had similar baseline vitamin concentrations; 1-month after supplement commencement, plasma concentrations became substantially higher in intervention participants; riboflavin by 0.77 SDS (95%CI 0.68-0.87), vitamin B6 1.07 (0.99-1.14), vitamin B12 0.55 (0.46-0.64) and vitamin D 0.51 (0.43-0.60), with the higher levels maintained during pregnancy and marked reduction in insufficiency/deficiency markers (lower homocysteine, hydroxykynurenine-ratio, methylmalonic acid) and the late pregnancy prevalence of vitamin D <50 nmol/L reduced from 35.1% to 8.5%. Plasma vitamin B12 was still higher in the intervention group 6-months post-delivery.

**Conclusion:** Significant proportions of preconception women have marginal or low status of folate, riboflavin, vitamin B12 and vitamin D, and many develop markers of vitamin B6 deficiency in late pregnancy. In the absence of supplementation, maternal plasma vitamin concentrations show differing longitudinal patterns from preconception to early and late pregnancy, suggesting plasma volume expansion does not wholly account for lower gestational concentrations. Preconception/pregnancy supplementation in amounts available in over-the-counter supplements substantially reduces the prevalence of deficiency/depletion markers before and during pregnancy, and a higher maternal plasma vitamin B12 was maintained during the recommended lactational period.

## Introduction

There is an increasing consensus that multiple micronutrient supplementation of pregnant women living in low-middle income countries is beneficial for pregnancy outcomes (1). In high income countries there are few large-scale trials of gestational micronutrient supplementation, resulting in less consensus about the need for individual or multiple micronutrient supplements to be taken. The principal exception relates to preconception and early pregnancy folic acid supplementation and fortification programs, underpinned by the landmark Medical Research Council trial (2). Vitamin D supplementation is also generally recommended, in part based on the recent MAVIDOS trial (3); MAVIDOS used a higher dose of vitamin D than is recommended in many settings (25 µg of cholecalciferol daily from early pregnancy until delivery, vs 10 µg daily recommended in countries such as the UK (4)), which reduced the incidence of infantile atopic eczema in the offspring (5) and improved measures of bone health in the children at age 4 years (6).

Recent evidence from animal studies demonstrates that maternal nutritional status prior to conception can have lasting effects on the offspring. This highlights a critical knowledge gap with regards to the importance of maternal micronutrient status before and during human pregnancy (7). Evidence from human studies supporting a role for preconception micronutrient status is largely observational, but does point to important implications for pregnancy outcomes and long-term offspring health (8). Examples include the associations of suboptimal vitamin B12 and B6 status with an increased risk of preterm birth (9), and of maternal preconception iodine deficiency with lower child IQ (10).

To date, maternal micronutrient status preconception has largely been inferred from pregnancy data, and truly longitudinal studies describing changes from preconception to early and late pregnancy and postpartum have not previously been conducted. The ongoing significant prevalence of micronutrient insufficiencies among adolescent girls and women of reproductive age in high income countries highlights the importance of documenting such changes (11). Moreover, lower pregnancy concentrations are often ascribed to plasma volume expansion (12, 13), without consideration of the longitudinal pattern or measurement of insufficiency/deficiency markers.

Human trials of micronutrient supplementation commencing before pregnancy remain relatively few in number; while they have not always shown benefits for maternal and offspring outcomes (14), a recent small trial did, however, report that maternal vitamin B12 supplementation from preconception until delivery improved offspring neurodevelopment at age 2 years (15). Development of evidence-based guidelines for micronutrient intake before/during pregnancy is additionally hindered by the paucity of randomized trial data on the change in maternal micronutrient status resulting from preconception and pregnancy supplementation using amounts available in over-the-counter supplements.

The NiPPeR trial (16) is a multicenter, double-blind, randomized controlled trial of a nutritional supplement containing micronutrients, myo-inositol and probiotics, whose primary outcome was the maintenance of euglycemia during pregnancy. The trial found no difference in gestational glycemia between study arms, but there was a significant reduction in preterm delivery, preterm pre-labor rupture of membranes and major post-partum hemorrhage with the intervention compared with controls, who received a standard micronutrient supplement (17). Longitudinal blood sampling in the NiPPeR trial provided an opportunity to determine the changes in maternal plasma concentrations of B-vitamins, vitamin D and insufficiency/deficiency markers (homocysteine, hydroxykynurenine-ratio, methylmalonic acid) from preconception, through early and late pregnancy and to 6 months postpartum (notably in the control group), and to examine the influence of maternal supplementation preconception and during pregnancy on mean concentrations and markers of low vitamin status.

## Subjects and Methods

### Trial study design

The study protocol has been previously published (16). Briefly, women planning a pregnancy were recruited from the community across 3 study sites in the UK, Singapore and New Zealand, between 2015-2017. Trial exclusion criteria were pregnancy/lactation at recruitment, assisted conception (apart from taking clomiphene or letrozole alone), serious food allergy, pre-existing diabetes mellitus, use of hormonal contraception or taking metformin, systemic steroids, anticonvulsants or treatment for HIV, Hepatitis B or C in the past month. With stratification by site and ethnicity to ensure balanced allocation, 1729 women were randomly assigned by an electronic database to receive intervention (n=870) or control (n=859) nutritional supplements from preconception until delivery. Maternal blood samples were collected from 870 intervention and 857 control women at preconception (at recruitment and one month after commencing supplementation), and then in early and late pregnancy, and six months post-delivery in those who became pregnant. Singleton pregnancies fulfilling the study criteria and reaching 28 weeks’ gestation with data on plasma vitamins at baseline and in late pregnancy were achieved in 580 women (Consort diagram shown in Supplementary Figure 1), with 512 followed up at 6 months post-delivery.

Intervention and control supplements with similar sensory characteristics and packaged as a powder in sachets were stored at 2-6°C until made up in 250 ml water and taken twice daily. Ingredients common to control and intervention formulations were folic acid 400 µg/day, iron 12 mg/day, calcium 150 mg/day, iodine 150 µg/day and β-carotene 720 µg/day; the intervention additionally included riboflavin 1.8 mg/day, vitamin B6 2.6 mg/day, vitamin B12 5.2 µg/day, vitamin D 10 µg/day, zinc 10 mg/day myo-inositol 4 g/day and probiotics (*Lactobacillus rhamnosus* and *Bifidobacterium animalis sp. lactis*). Quantities were either UK recommended daily allowances for pregnant women (vitamin D, zinc, folic acid, iodine), minimal amounts for micronutrients linked with potential detrimental effects at higher doses (iron, β-carotene, calcium), or amounts enhanced above those typical in over-the-counter products (vitamins B6, B12, riboflavin) or used in previous trials (myo-inositol, probiotics) (18, 19). Supplements were consumed from preconception following randomization until delivery of the baby. Participants and all study personnel remained blinded to treatment allocation until all pregnancy, delivery and neonatal data had been collected, and analysis of the primary outcome completed. At enrolment, sociodemographic characteristics, menstrual, obstetric and health histories and lifestyle habits were collected via interviewer-administered questionnaires. Weight and height were measured to derive body mass index (BMI). Adherence to the trial formulation ascertained by sachet counting was similar in the control and intervention groups; overall, 80.6% had 80-100% adherence averaged from recruitment to delivery, 15.8% had 60-80% adherence, and only 3.1% adherence below 60%. Women were recommended to refrain from taking other supplements unless advised by their healthcare practitioner (e.g. significant iron deficiency anemia).

The trial was approved by the United Kingdom, Singapore and New Zealand research ethics services (Southampton: Health Research Authority NRES Committee South Central Research Ethics Committee, reference 15/SC/0142l; Singapore: the National Healthcare Group Domain Specific Review Board, reference 2015/00205; New Zealand: the Health and Disability Ethics Committee, reference 15/NTA/21). The relevant regulatory authorities confirmed that the formulation was not an investigational medicinal product. All participants gave written informed consent. Trial oversight and monitoring were provided by an independent data and safety monitoring committee. This trial was prospectively registered at ClinicalTrials.gov NCT02509988, UTN U1111-1171-8056.

### Vitamin, vitamer and metabolite measurements and status markers

EDTA plasma was obtained by centrifugation of peripheral venous blood samples at 1600 g at 4°C for 10 minutes and stored at -80°C for later analyses. Aliquots were available for measurement of plasma vitamins and related vitamers/metabolites at the following five time-points: preconception at recruitment (n=1721) and 1 month after commencing intervention (n=1454), in early (7-11 weeks’ gestation, n=634) and late pregnancy (around 28 weeks’ gestation, n=580), and post-delivery, 6 months after discontinuation of supplementation (n=512). Using a targeted method based on liquid chromatography-tandem mass spectrometry (Bevital, Bergen, Norway)(20), we measured plasma concentrations of vitamins present in the control and intervention groups, related vitamers and metabolites that reflect vitamin status: homocysteine (reflecting 1-carbon status and an indicator of folate and B-vitamin deficiency), riboflavin, flavin mononucleotide (reflecting riboflavin status), pyridoxal 5-phosphate (vitamin B6), 3-hydroxykynurenine (HK), kynurenic acid (KA), anthranilic acid (AA), 3-xanthurenic acid (XA), hydroxyanthranilic acid (HAA), cystathionine, cysteine, methylmalonic acid and 25-hydroxyvitamin D3. Plasma folate and cobalamin (vitamin B12) were measured by microbiological assay, using a microtiter plate format on a robotic workstation employing a chloramphenicol-resistant strain of Lactobacillus casei (folate) and a colistin sulphate-resistant strain of Lactobacillus leichmannii (cobalamin)(Bevital, Bergen, Norway). Detailed quality control data for all analytes has been described previously, documenting coefficients of variation <10% (21). Values below the assay limit of detection were set to half the limit of detection value. Any hemolysis was visually graded from zero to 4+; anthranilic acid, 3-hydroxyanthranilic acid and 3-hydroxykynurenine values were set to missing for samples with a hemolysis score ≥2 (approximately equivalent to Hb 250 mg/dL), and those for folate set to missing for samples with 4+ hemolysis (approximately equivalent to Hb 1000 mg/dL). Each analyte was checked for outliers (both statistically and clinically), and implausible values were set to missing.

As functional markers of vitamin B6 status, we calculated the plasma 3´-hydroxykynurenine ratio (HK ratio [HK:KA+AA+XA+HAA], a higher ratio reflecting effects of vitamin B6 insufficiency on tryptophan catabolism), along with the cystathionine/cysteine ratio (a higher ratio reflecting effects of vitamin B6 insufficiency on transsulfuration pathway regulation) (22). There is inconsistency in the literature regarding thresholds for vitamin deficiency or insufficiency markers based on plasma measurements: for this study we used plasma folate 13.6 nmol/L to define “marginal folate status” (23); homocysteine ≥15 µmol/L to define “high plasma homocysteine” (24); riboflavin <5 nmol/L (the lower limit of the reference range for the laboratory) and ≤26.5 nmol/L (the change-point of plasma riboflavin with erythrocyte glutathione reductase activation coefficient (25)) to define low riboflavin and “marginal riboflavin status”, respectively; pyridoxal 5-phosphate <20 nmol/L to define low B6 status; cobalamin <148 and <221 pmol/L to define B12 “deficiency” and “depletion”, respectively (26); and plasma 25-hydroxyvitamin D3 <50 and <75 nmol/L to define “deficiency” and “insufficiency”, respectively (27).

### Statistics

The sample size was based on the trial primary outcome of gestational glycemia, as described previously (17). Following log_e_ (natural logarithm) transformation where necessary, standardization was applied to all values of each analyte (i.e. ∼4902 samples) to derive standard deviation scores (SDS) before being split into separate variables by time-point/visit; the advantage of this approach is that for each analyte the effect sizes are comparable between time-points.

For each of the vitamins and related vitamers and metabolites analyses focused on: i) describing the longitudinal changes in maternal concentrations from preconception through pregnancy and to 6 months post-delivery in the control group, ii) the pattern of longitudinal change in the intervention group, and iii) differences in concentration and in sufficiency markers between the control and intervention groups at different time-points, including differences in proportions analyzed using chi-squared tests. Longitudinal plots are shown as means and 95% confidence intervals (CI) in SDS by intervention group on y-axis and time-point/visit on x-axis, with statistical significance considered when the 95%CI did not overlap (two-tailed P<0.05). Differences given in the text are statistically significant unless otherwise stated. Plots and differences in SDS between time-points are based on all available data at each time-point unless otherwise specified. Sensitivity analyses were used to determine whether differences between time-points reflected differences between those who did and did not become pregnant (resulting in differing numbers of participants with preconception, pregnancy and post-delivery measurements), and linear and logistic regression analyses were used to ascertain whether the patterns of longitudinal and control vs intervention group differences were robust to adjustment for site and ethnicity (trial randomization stratification factors) and parity (not fully balanced across control and intervention groups and potentially influential on status measurements). Analyses were performed using Stata software v15.1 (StataCorp, College Station, TX, USA).

## Results

Characteristics of the participants with preconception data (n=1727), and of those who had baseline analyte data and were followed up at late pregnancy (n=580) are shown in Table 1. Characteristics were balanced across the control and intervention groups, except for somewhat more nulliparity among controls; characteristics for those followed up post-delivery (n=512) were similar to those with preconception data.

**Table 1.** Participant characteristics.

|  |  | Preconception |  | Baseline and late pregnancy<sup>+</sup> |  |
| --- | --- | --- | --- | --- | --- |
| Characteristic |  | Control (n=857) | Intervention (n=870) | Control (n=287) | Intervention (n=293) |
| *Sociodemographic* |  |  |  |  |  |
| Study site | UK | 229 (26.7%) | 232 (26.7%) | 92 (32.1%) | 97 (33.1%) |
|  | Singapore | 328 (38.3%) | 332 (38.2%) | 82 (28.6%) | 84 (28.7%) |
|  | New Zealand | 300 (35.0%) | 306 (35.2%) | 113 (39.4%) | 112 (38.2%) |
| Age, years | Mean (SD) | 30.6 (3.7) | 30.6 (3.7) | 30.2 (3.3) | 30.5 (3.4) |
| Ethnicity | White | 396 (46.2%) | 413 (47.5%) | 165 (57.5%) | 179 (61.1%) |
|  | Chinese | 220 (25.7%) | 238 (27.4%) | 73 (25.4%) | 72 (24.6%) |
|  | South Asian | 60 (7.0%) | 61 (7.0%) | 15 (5.2%) | 15 (5.1%) |
|  | Malay | 85 (9.9%) | 80 (9.2%) | 12 (4.2%) | 11 (3.8%) |
|  | Other | 96 (11.2%) | 78 (9.0%) | 22 (7.7%) | 16 (5.5%) |
| Household income<sup>#</sup> | Low income | 74 (9.3%) | 76 (9.4%) | 11 (4.0%) | 12 (4.3%) |
|  | Middle income | 344 (43.2 %) | 354 (43.7%) | 114 (41.2%) | 114 (40.9%) |
|  | High income | 378 (47.5%) | 380 (46.9%) | 152 (54.9%) | 153 (54.8%) |
| *Gynecological* |  |  |  |  |  |
| Parity | Nulliparous | 593 (69.2%) | 578 (66.4%) | 196 (68.3%) | 169 (57.7%) |
|  | Parous | 264 (30.8%) | 292 (33.6%) | 91 (31.7%) | 124 (42.3%) |
| *Lifestyle* |  |  |  |  |  |
| Alcohol intake (per week) | None | 271 (31.6%) | 265 (30.5%) | 61 (21.3%) | 65 (22.2%) |
|  | >0 to ≤2.5 units | 312 (36.4%) | 301 (34.6%) | 115 (40.1%) | 109 (37.2%) |
|  | >2.5 units | 274 (32.0%) | 304 (34.9%) | 111 (38.7%) | 119 (40.6%) |
| Smoking status | Never | 664 (77.7%) | 662 (76.3%) | 225 (78.7%) | 237 (80.9%) |
|  | Previous | 131 (15.3%) | 136 (15.7%) | 49 (17.1%) | 44 (15.0%) |
|  | Active | 60 (7.0%) | 70 (8.1%) | 12 (4.2%) | 12 (4.1%) |
| Instances of moderate /vigorous physical activity in past 7 days | Median (IQR) | 3 (2, 5) | 3 (1, 5) | 4 (2, 6) | 3 (2, 5) |
| Body mass index (BMI) category<sup>##</sup> | Not overweight or obese | 400 (46.7%) | 432 (49.8%) | 158 (55.1%) | 163 (55.8%) |
|  | Overweight | 240 (28.0%) | 224 (25.8%) | 69 (24.0%) | 89 (30.5%) |
|  | Obese | 216 (25.2%) | 212 (24.4%) | 60 (20.9%) | 40 (13.7%) |
| Days between preconception baseline and post-supplementation sampling | Median (IQR) | 28 (23, 31) | 28 (23, 32) | 27 (22, 31) | 28 (23, 31) |

Data presented as number (%) unless otherwise stated. Sample sizes do not always equal to 857/251 for control group and 870/261 for intervention group due to missing values; <sup>+</sup> based on all available data for individuals with any analyte at both baseline and late pregnancy <sup>#</sup>Low 1<sup>st</sup>-3<sup>rd</sup> decile, Middle 4<sup>th</sup>-7<sup>th</sup> decile, High 8<sup>th</sup>-10<sup>th</sup> decile; <sup>##</sup>BMI categories - Not overweight or obese, Overweight and Obese <25, 25-<30 and ≥30 kg/m<sup>2</sup>, respectively Abbreviations: IQR, interquartile range; SD, standard deviation

Table 2 shows median and interquartile range values for the maternal plasma vitamins and related vitamers/metabolites in their original units at each time-point for each of the control and intervention groups, and Table 3 the percentages of participants below vitamin and vitamer deficiency/insufficiency marker thresholds according to time-point and control/intervention group. The longitudinal changes in maternal concentrations expressed in SDS and proportions with marginal or low status, depletion, deficiency and insufficiency are shown graphically in Figures 1-5. Among all participants at recruitment preconception, significant proportions had marginal or low plasma status for folate (29.2% <13.6 nmol/L), riboflavin (7.5% <5 nmol/L, 82.0% ≤26.5 nmol/L), vitamin B12 (9.1% <221 pmol/L) and vitamin D (48.7% <50 nmol/L); only 1.3% had a low pyridoxal 5-phosphate (<20 nmol/L).

**Table 2:** Median (IQR) values in original units according to control or intervention group at each time-point.

|  | Preconception baseline |  | Preconception 1-month post supplementation |  | Early pregnancy (7-11 weeks gestation) |  | Late pregnancy (28 weeks gestation) |  | 6 months post-delivery |  |
| --- | --- | --- | --- | --- | --- | --- | --- | --- | --- | --- |
|  | Controls | Intervention | Controls | Intervention | Controls | Intervention | Controls | Intervention | Controls | Intervention |
| Folate (nmol/L) | 22.5<br>(12.7, 41.3) | 20.5<br>(12.1, 41.3) | 41.9<br>(29.3, 55.0) | 40.3<br>(29.0, 52.8) | 49.8<br>(40.5, 60.0) | 52.2<br>(43.2, 63.0) | 41.5<br>(30.6, 51.6) | 44.1<br>(34.3, 54.5) | 19.2<br>(11.9, 32.7) | 18.9<br>(12.3, 32.2) |
| Homocysteine (µmol/L) | 7.0<br>(6.1, 8.4) | 7.1<br>(6.1, 8.5) | 6.9<br>(6.0, 8.0) | 6.4<br>(5.6, 7.4) | 5.5<br>(4.8, 6.4) | 4.9<br>(4.2, 5.6) | 4.6<br>(3.9, 5.2) | 3.9<br>(3.5, 4.6) | 7.4<br>(6.4, 9.0) | 7.2<br>(6.1, 8.5) |
| Riboflavin (nmol/L) | 12.7<br>(7.7, 21.7) | 12.0<br>(7.6, 21.2) | 12.7<br>(8.3, 22.3) | 24.7<br>(16.7, 36.8) | 10.7<br>(6.8, 18.0) | 20.0<br>(13.0, 30.3) | 10.3<br>(6.6, 15.1) | 16.9<br>(12.2, 23.3) | 12.9<br>(8.8, 21.1) | 12.7<br>(8.4, 22.6) |
| FMN (nmol/L) | 14.4<br>(11.5, 18.7) | 14.6<br>(11.9, 18.7) | 15.6<br>(12.4, 20.1) | 18.4<br>(15.0, 23.4) | 14.5<br>(11.4, 18.2) | 17.2<br>(14.3, 21.6) | 10.1<br>(8.4, 12.0) | 11.5<br>(9.8, 13.4) | 14.2<br>(11.8, 17.0) | 14.4<br>(11.2, 18.7) |
| Pyridoxal 5-phosphate (nmol/L) | 55.8<br>(40.3, 88.4) | 57.1<br>(41.1, 84.3) | 56.9<br>(43.0, 80.1) | 140.0<br>(102.0, 184.0) | 47.6<br>(34.3, 64.1) | 106.0<br>(76.0, 139.0) | 19.1<br>(14.7, 28.1) | 41.9<br>(30.3, 56.4) | 56.3<br>(40.0, 88.2) | 56.1<br>(40.5, 86.6) |
| HK ratio (no units) | 0.35<br>(0.29, 0.43) | 0.36<br>(0.29, 0.42) | 0.35<br>(0.29, 0.42) | 0.29<br>(0.24, 0.35) | 0.35<br>(0.28, 0.43) | 0.29<br>(0.24, 0.33) | 0.51<br>(0.41, 0.60) | 0.44<br>(0.37, 0.54) | 0.40<br>(0.32, 0.49) | 0.39<br>(0.32, 0.47) |
| Cobalamin (pmol/L) | 355.2<br>(283.1, 434.1) | 350.9<br>(278.8, 437.7) | 358.9<br>(284.0, 449.3) | 438.2<br>(353.3, 552.1) | 307.4<br>(240.2, 384.0) | 411.1<br>(332.0, 509.3) | 214.6<br>(177.3, 270.0) | 298.7<br>(247.9, 379.5) | 340.4<br>(264.5, 437.8) | 385.6<br>(312.3, 465.7) |
| MMA (µmol/L) | 0.13<br>(0.11, 0.17) | 0.13<br>(0.11, 0.17) | 0.15<br>(0.12, 0.19) | 0.15<br>(0.12, 0.19) | 0.12<br>(0.10, 0.15) | 0.11<br>(0.10, 0.14) | 0.14<br>(0.11, 0.20) | 0.12<br>(0.10, 0.15) | 0.15<br>(0.12, 0.20) | 0.14<br>(0.12, 0.17) |
| Vitamin D3 (nmol/L) | 51.0<br>(36.0, 65.3) | 50.1<br>(36.1, 65.8) | 51.7<br>(37.0, 65.5) | 61.1<br>(51.1, 73.3) | 53.3<br>(41.8, 69.5) | 69.6<br>(57.3, 81.2) | 62.9<br>(41.0, 87.5) | 92.8<br>(73.0, 106.9) | 62.5<br>(45.3, 77.6) | 64.6<br>(48.5, 77.6) |

HK ratio – 3'-hydroxykynurenine ratio, FMN – flavin mononucleotide, MMA – methylmalonic acid

**Table 3:**
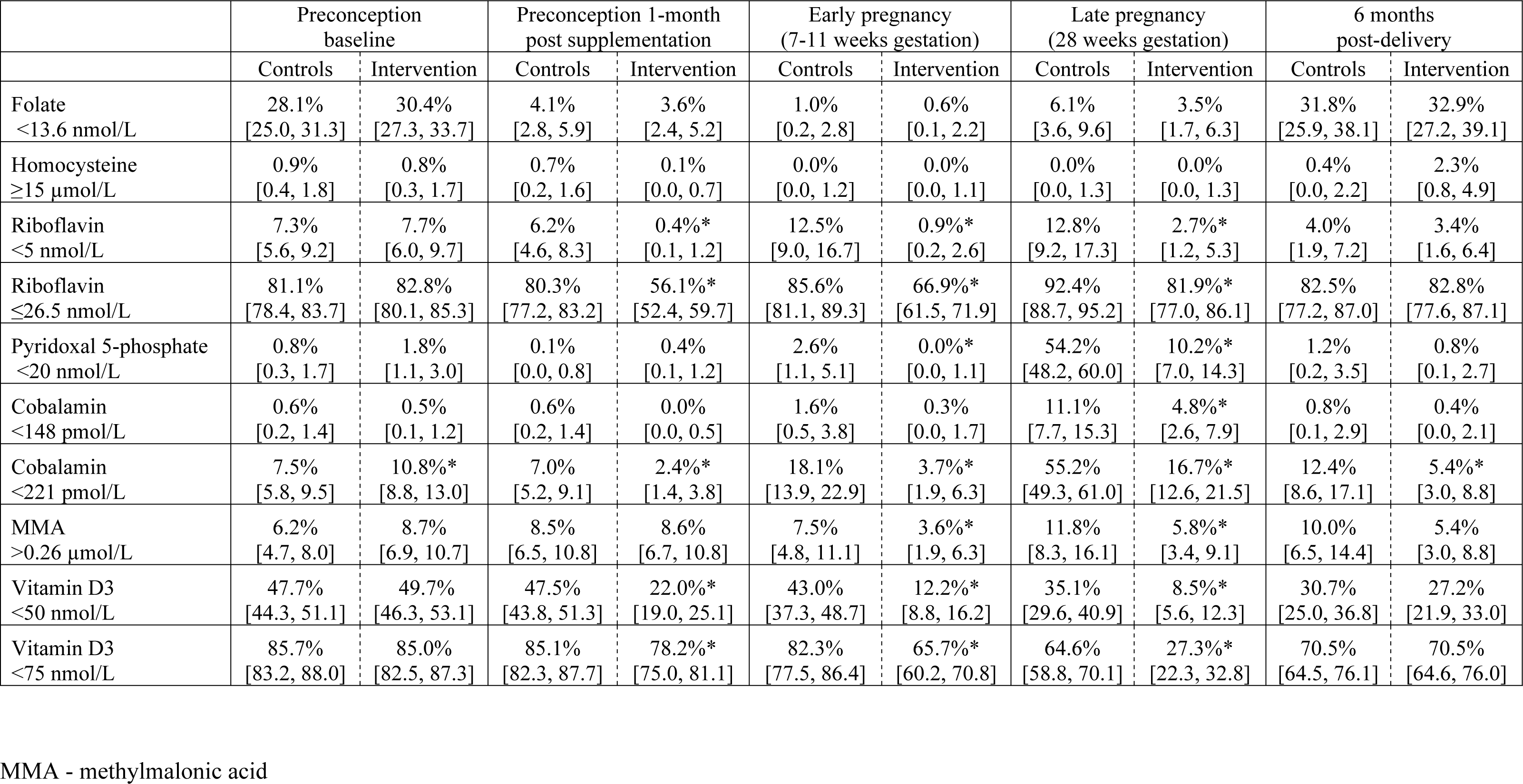
Percentages of participants meeting vitamin/vitamer deficiency/insufficiency thresholds according to time-point and control/ntervention group. Values in parentheses are 95% confidence intervals. *Chi-squared P<0.05 for intervention vs control difference at that time point

**Figure 1:**
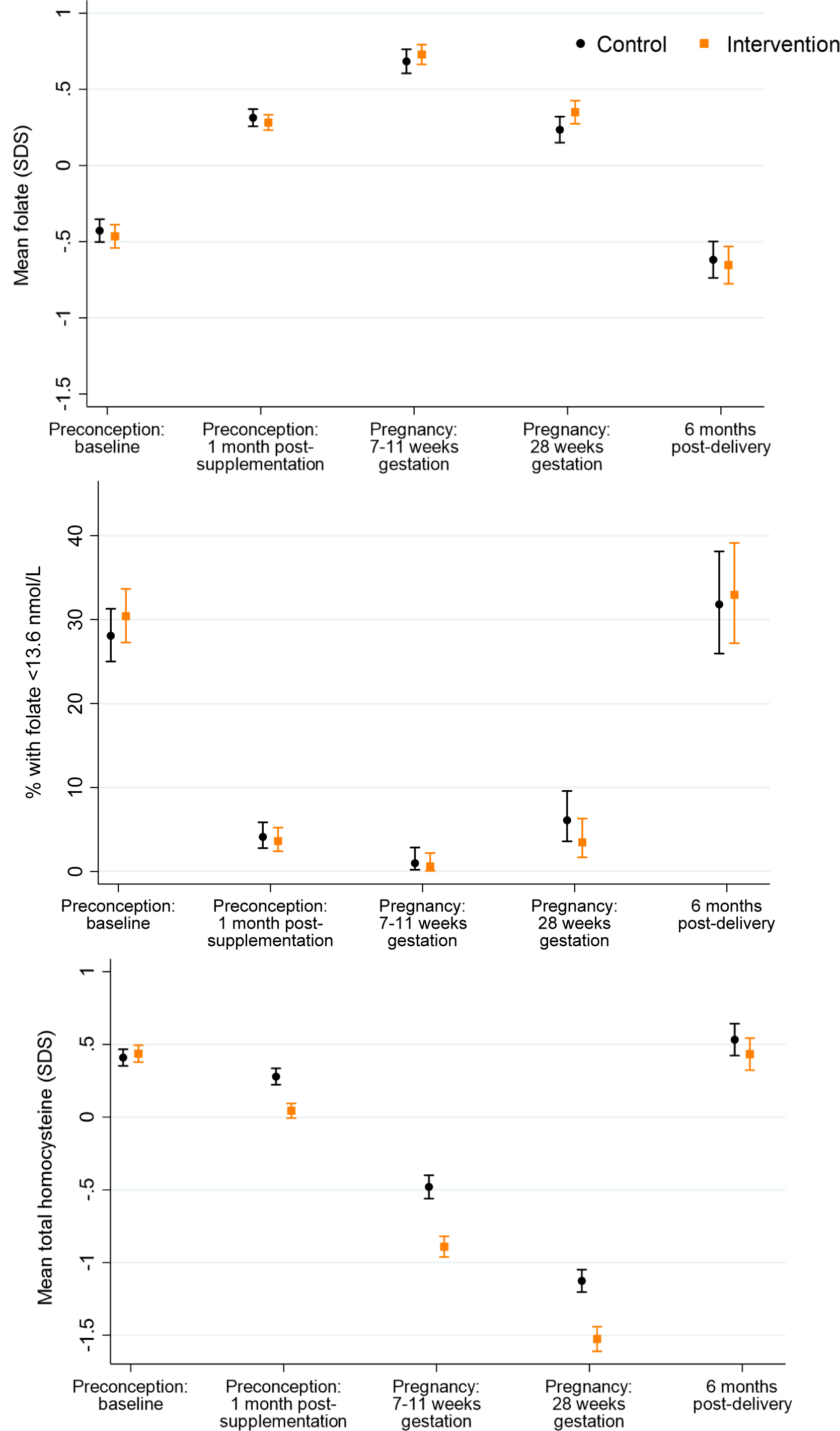
Plasma folate, marginal folate status and plasma homocysteine according to time-point and control vs intervention group.

### Plasma folate and homocysteine

In the control group (who received 400 µg/day folic acid, as in the intervention group), from preconception baseline to 1-month after supplementation commencement mean maternal plasma folate increased by 0.74 SDS, followed by a further 0.37 SDS increase in early pregnancy, and then declined 0.45 SDS from early to late pregnancy and 0.85 SDS from late pregnancy to 6-months post-delivery, resulting in a post-delivery concentration lower than at preconception baseline (Figure 1a); marginal folate status (<13.6 nmol/L) was present in 28.1% of control group participants at preconception baseline, falling to 4.1% 1-month after supplementation commencement and 1.0% in early pregnancy, before rising to 6.1% in late pregnancy and 31.8% 6-months post-delivery (Figure 1b). The intervention group showed a similar longitudinal pattern for plasma folate.

Plasma homocysteine concentrations, an indicator of folate and B-vitamin deficiency, in the control group decreased by 0.13 SDS from preconception baseline to 1-month after supplementation commencement, followed by further 0.76 and 0.65 SDS falls in early and late pregnancy, respectively, and then a 1.66 SDS increase 6-months post-delivery, resulting in post-delivery concentration marginally higher than at recruitment (Figure 1c); a high plasma homocysteine ≥15 µmol/L was present in 0.9% of participants at preconception baseline, falling to 0% in early pregnancy and late pregnancy, and 0.4% 6 months post-delivery. The intervention group showed a similar longitudinal pattern, but plasma homocysteine concentrations were 0.24, 0.41 and 0.40 SDS lower than in the control group 1-month after supplementation commencement and in early and late pregnancy, respectively.

### Plasma riboflavin and flavin mononucleotide

Among control group participants (taking a supplement without riboflavin), as expected plasma riboflavin was similar at preconception baseline and 1-month after supplementation commencement, then decreased by 0.24 SDS in early pregnancy, with a further 0.17 SDS decrease in late pregnancy, followed by an increase of 0.43 SDS at 6-months post-delivery, returning to levels similar to those at preconception (Figure 2a). Marginal riboflavin status (≤26.5 nmol/L) was present in 81.1% of control participants at preconception baseline and 80.3% 1-month after supplementation commencement, increasing to 85.6% and 92.4% in early and late pregnancy, respectively, before returning to 82.5% 6-months post-delivery (Supplementary Figure 2). Low riboflavin status (<5 nmol/L) in the control group showed a similar longitudinal pattern at the expected lower prevalence (7.3%, 6.2%, 12.5%, 12.8% and 4.0%, across the five time-points)(Figure 2b). Compared with the control group, plasma riboflavin concentrations in the intervention group 1-month after supplementation commencement, in early pregnancy and in late pregnancy were higher (by 0.77, 0.76 and 0.65 SDS, respectively), with lower prevalences of marginal (56.1%, 66.9% and 81.9%, respectively) and low (0.4%, 0.9% and 2.7%, respectively) riboflavin status.

**Figure 2:**
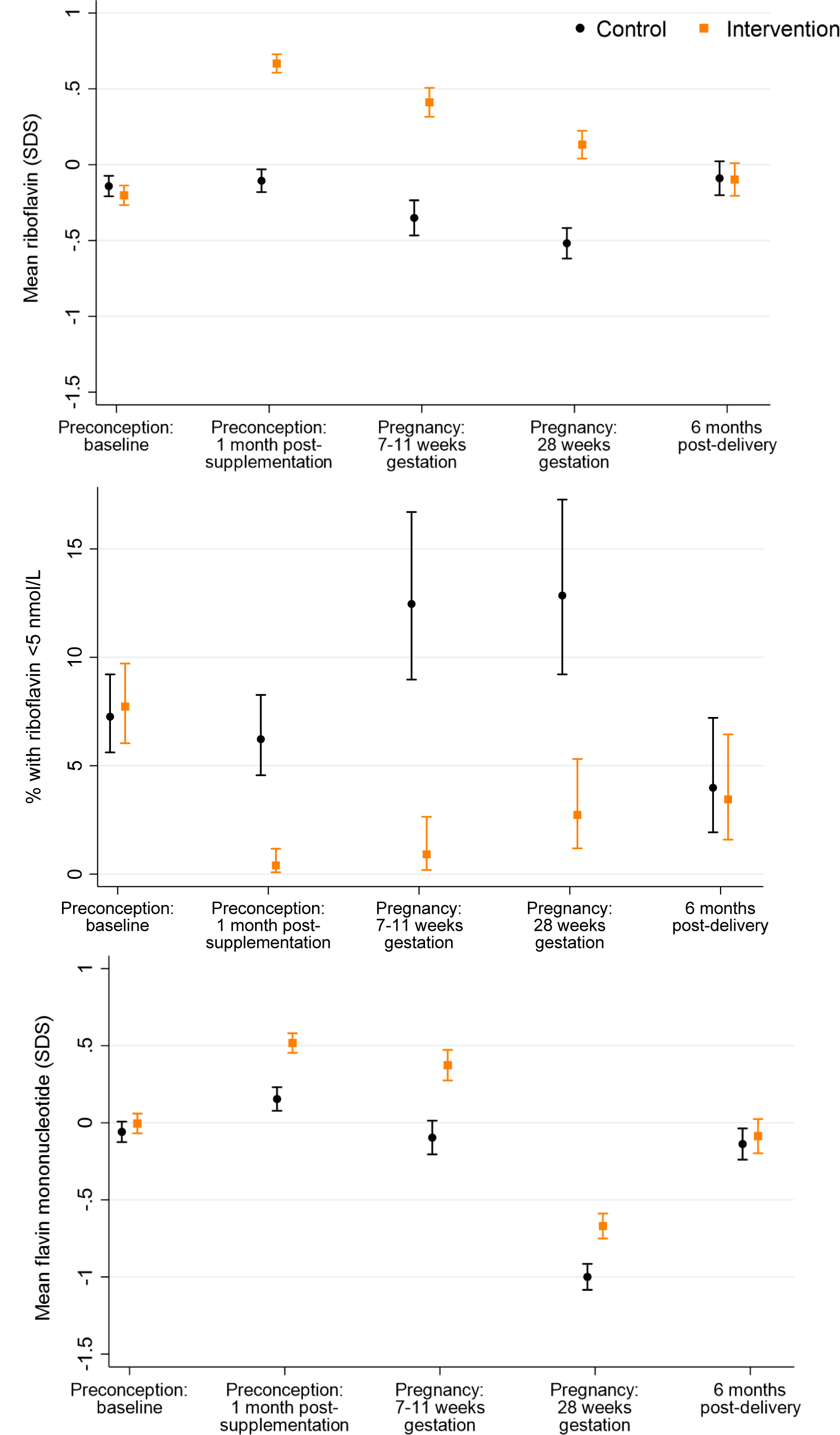
Plasma riboflavin, low riboflavin and plasma flavin mononucleotide according to time-point and control vs intervention group.

Plasma flavin mononucleotide, a marker of riboflavin sufficiency, in the control group showed a similar longitudinal pattern to plasma riboflavin but increased by 0.21 SDS from preconception baseline to 1-month after supplementation commencement, before showing a similar decline during pregnancy (Figure 2c). Plasma flavin mononucleotide concentrations were 0.36, 0.47 and 0.33 SDS higher in the intervention vs the control group 1-month after supplementation commencement, in early pregnancy and in late pregnancy, respectively.

### Plasma vitamin B6 and B6 insufficiency markers

In the control group (taking a supplement without vitamin B6), plasma pyridoxal 5-phosphate was similar at preconception baseline and 1-month after supplementation commencement, then decreased by 0.25 SDS between 1-month after supplementation commencement and early pregnancy, before a substantial further decrease of 1.11 SDS from early pregnancy to late pregnancy, followed by a return to preconception concentrations 6-months post-delivery (Figure 3a); preconception, low pyridoxal 5-phosphate (<20 nmol/L) was present in 0.8% of participants at baseline and 0.1% 1-month after supplementation commencement, increasing to 2.6% in early pregnancy and a steep rise to 54.2% in late pregnancy, before returning to 1.2% 6-months post-delivery (Figure 3b). Compared with the control group, the intervention group had higher plasma pyridoxal 5-phosphate concentrations at 1-month after supplementation commencement, and in early and late pregnancy, by 1.07, 0.95 and 0.84 SDS, respectively.

**Fig. 3:**
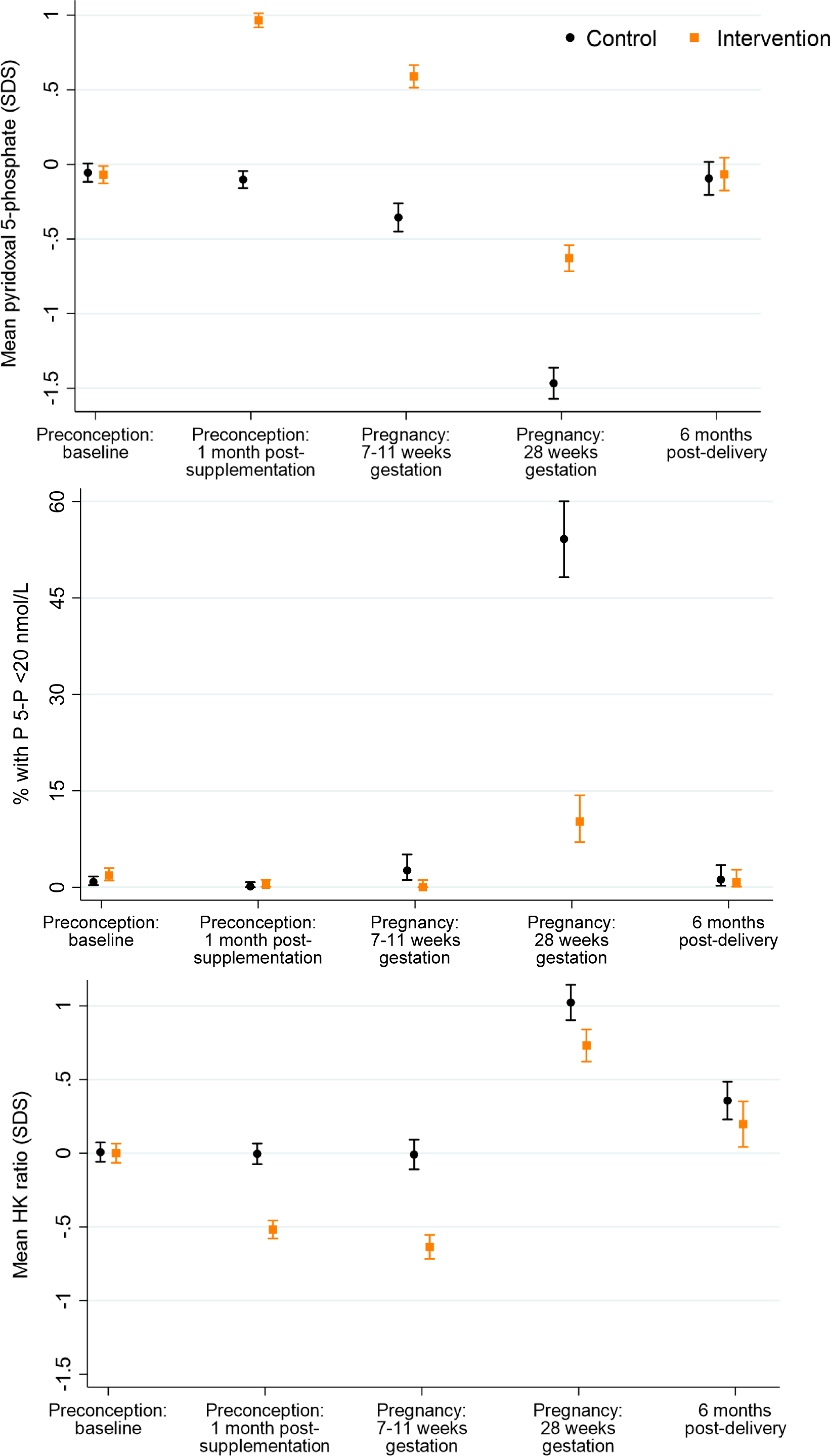
Plasma pyridoxal 5-phosphate (P 5-P), low pyridoxal 5-phosphate and plasma 3-hydroxyknurenine (HK) ratio according to time-point and control vs intervention group.

Among control participants, the plasma HK ratio was unchanged from the preconception baseline through 1-month after supplementation commencement and in early pregnancy, but then rose sharply by 1.03 SDS between early and late pregnancy, reflecting reduced activity of vitamin B6 dependent pathways; between late pregnancy and 6-months post-delivery the HK ratio fell by 0.67 SDS, to a level that was higher than the preconception baseline (Figure 3c). Compared with the control group, in the intervention group plasma HK ratio was lower at 1-month after supplementation commencement, in early pregnancy and in late pregnancy, by 0.51, 0.63 and 0.29 SDS, respectively. In the control group, the plasma cystathionine/cysteine ratio increased from preconception baseline to 1-month after supplementation commencement, decreasing to a level slightly lower than baseline in early pregnancy, followed by a 1.36 SDS rise between early and late pregnancy, and then a fall 6-months post-delivery to a level higher than the preconception baseline (Supplementary Figure 2). Similar to the HK ratio, compared with the control group, the plasma cystathionine/cysteine ratio was lower in the intervention group 1-month after supplementation commencement, in early pregnancy and in late pregnancy, by 0.35, 0.37 and 0.23 SDS, respectively.

### Plasma vitamin B12 and methylmalonic acid

In the control group (taking a supplement without vitamin B12), plasma cobalamin was similar at preconception baseline and 1-month after supplementation commencement, then decreased by 0.41 SDS in early pregnancy, further decreased by 0.88 SDS in late pregnancy, then returning to baseline preconception concentrations 6-months post-delivery (Figure 4a); preconception, vitamin B12 deficiency (<148 pmol/L) was present in 0.6% of participants at baseline and 1-month after supplementation commencement, increasing to 1.6% and 11.1% in early and late pregnancy, respectively, before returning to 0.8% 6-months post-delivery (Supplementary Figure 4). Vitamin B12 depletion (<221 pmol/L) in the control group showed a similar longitudinal pattern at the expected higher prevalence (7.5%, 7.0%, 18.1%, 55.2% and 12.4%, across the five time-points) (Figure 4b). Compared with the control group, in the intervention group plasma cobalamin concentrations 1-month after supplementation commencement, in early pregnancy and in late pregnancy were higher (by 0.55, 0.75 and 0.79 SDS, respectively), with lower prevalences of vitamin B12 deficiency (0%, 0.3% and 4.8%, respectively), and of vitamin B12 depletion (2.4%, 3.7% and 16.7%, respectively). Notably, plasma vitamin B12 was 0.3 SDS higher in the intervention group than the control group 6-months post-delivery, with a lower prevalence of vitamin B12 depletion (5.4% vs 12.4%).

**Figure 4:**
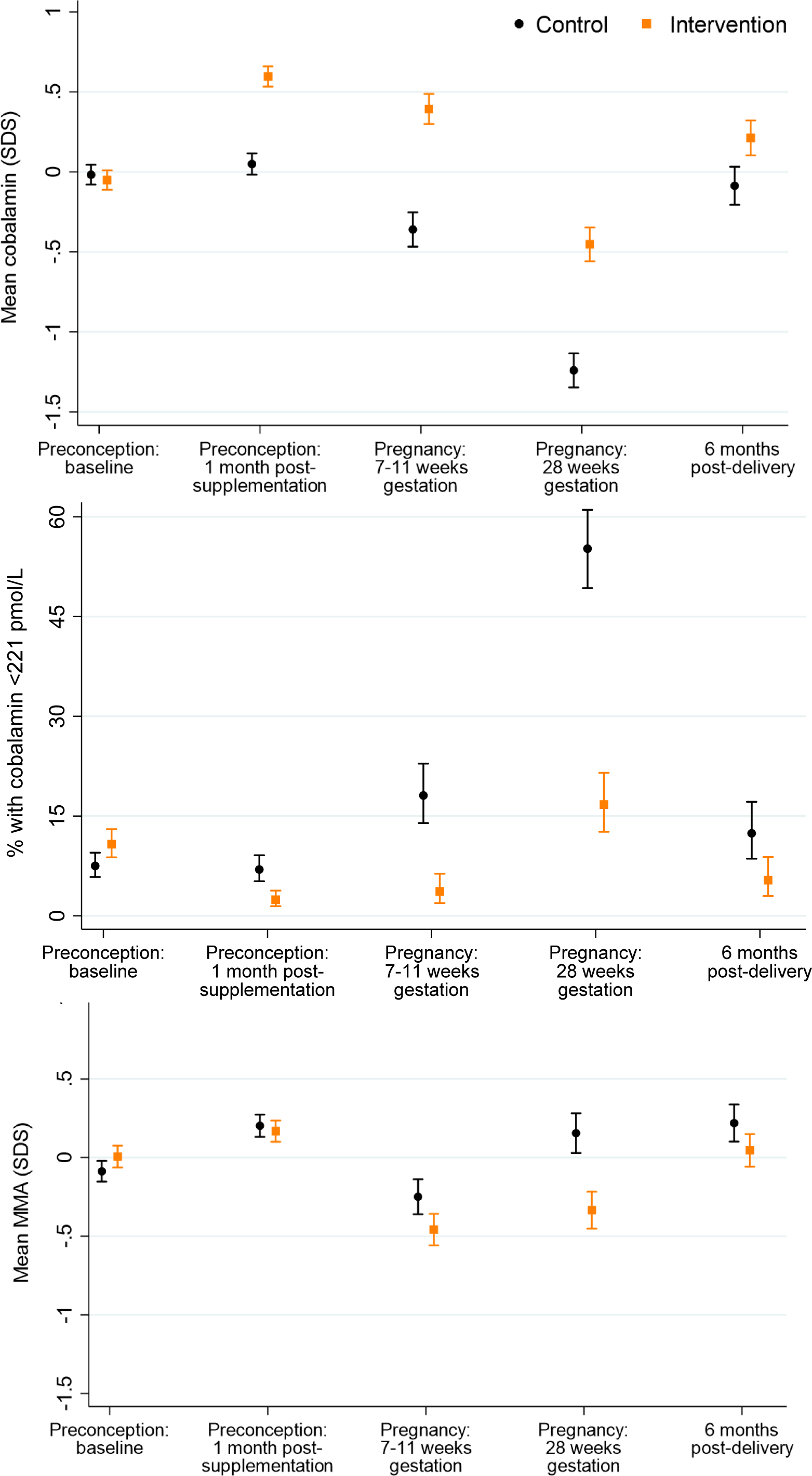
Plasma cobalamin, cobalamin depletion and plasma methylmalonic acid (MMA) according to time-point and control vs intervention group.

Plasma methylmalonic acid, a metabolic indicator of vitamin B12 insufficiency, in the control group increased by 0.29 SDS from preconception baseline to 1-month after supplementation commencement, then decreasing by 0.45 SDS in early pregnancy before increasing 0.39 SDS in late pregnancy and staying at a similar level 6-months post-delivery, 0.31 SDS above the original preconception baseline value (Figure 2c). Plasma methylmalonic acid concentrations in the intervention group were similar to those in the control group 1-month after supplementation commencement, but 0.21 and 0.49 SDS lower in early pregnancy and in late pregnancy, respectively; 6-months post-delivery there was a 0.17 SDS lower concentration in the intervention group (P=0.05).

### Plasma 25-hydroxyvitamin D

In the control group, plasma 25-hydroxyvitamin D changed little from preconception baseline to 1-month after supplementation commencement and early pregnancy, then increased by 0.28 SDS between early and late pregnancy, remaining similar to late pregnancy concentrations 6-months post-delivery (Figure 5a). Vitamin D deficiency <50 nmol/L in the control group showed a similar longitudinal pattern, with high prevalences at preconception baseline, 1-month after supplementation commencement and in early pregnancy (47.7%, 47.5% and 43.0%, respectively), and rather lower prevalences in late pregnancy (35.1%) and 6-months post-delivery (30.7%)(Figure 5b); vitamin D insufficiency <75 nmol/L showed a similar longitudinal pattern at high prevalence levels (85.7%, 85.1%, 82.3%, 64.6%, and 70.5%, across the five time-points)(Figure 5c). Compared with the control group, in the intervention group plasma 25-hydroxyvitamin D concentrations 1-month after supplementation commencement, in early pregnancy and in late pregnancy were higher (by 0.51, 0.63 and 0.89 SDS, respectively), with prevalences of vitamin D deficiency (22.0%, 12.2% and 8.5%, respectively) and insufficiency (78.2%, 65.7% and 27.3%, respectively) that were substantially lower than the control group; 6-months post-delivery plasma 25-hydroxyvitamin D concentrations were similar in the intervention and control groups (Figures 5b, 5c).

**Figure 5:**
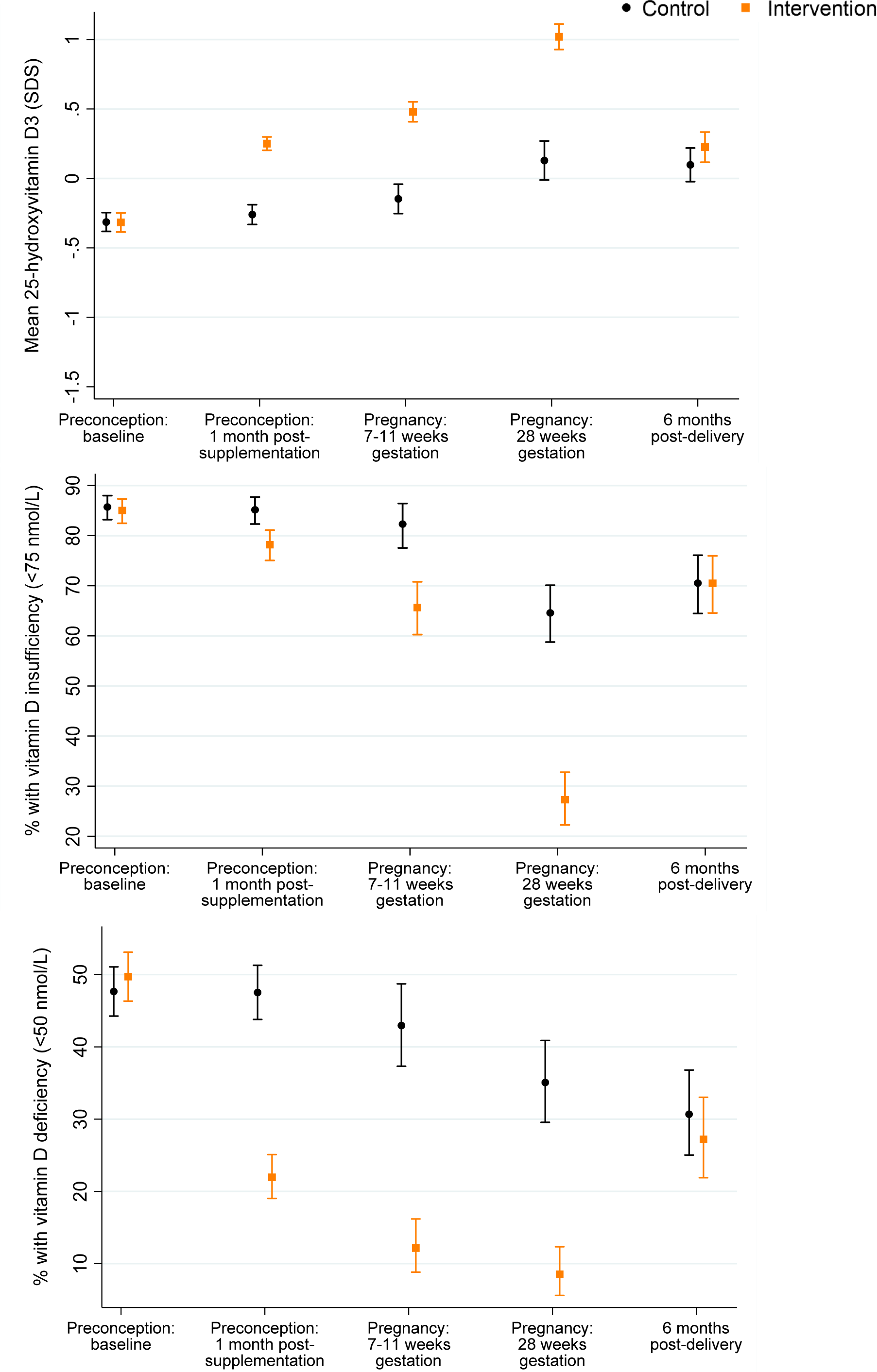
Plasma 25-hydroxyvitamin D, vitamin deficiency and insufficiency according to time-point and control vs intervention group.

### Sensitivity analyses

In sensitivity analyses of participants with complete preconception, pregnancy and post-delivery data the patterns described above for each of the vitamins and vitamers/metabolites were confirmed as little different, with no substantive differences from the analyses of all participants with measurements at any time-point (Supplementary Table 1). Further sensitivity analyses showed that the patterns of longitudinal and control vs intervention group differences were robust to adjustment for site, ethnicity and parity.

## Discussion

Among UK, Singapore and New Zealand women attempting to become pregnant in this multi-center randomized controlled trial, significant proportions had marginal or low status of folate, riboflavin, vitamin B12 and vitamin D at recruitment preconception, and a high proportion developed markers of functional vitamin B6 deficiency in late pregnancy despite a small proportion having a low vitamin B6 status preconception. Plasma concentrations for the above vitamins, related vitamers and metabolic insufficiency markers showed differing patterns of change from preconception to pregnancy and 6-months post-delivery. A formulation enriched with micronutrients in amounts available in over-the-counter supplements commenced preconception and continued throughout pregnancy substantially increased plasma folate, riboflavin and vitamins B6, B12 and D concentrations and reduced the prevalence of their deficiency/depletion markers before and during pregnancy, with a persisting benefit of higher plasma vitamin B12 to 6-months post-delivery.

As routinely recommended for preconception and pregnancy, supplementation with 400 µg folic acid daily in both the control and intervention groups raised plasma folate by >1 SDS from baseline to early pregnancy and reduced the prevalence of marginal folate status from 28.1% at baseline to 1.0% in early pregnancy. Our findings show that even with 400 µg, the lowest commonly marketed dose, there are appreciable improvements in folate status after 1 month and continued improvement into pregnancy. Plasma homocysteine in the control group fell substantially from preconception baseline to early pregnancy and again from early to late pregnancy, with greater falls in plasma homocysteine in the intervention group taking a supplement containing other micronutrients. While some have proposed that vitamin B6, riboflavin and zinc might reduce homocysteine, there are data pointing away from this (28, 29). One previous review concluded that dietary folic acid reduced homocysteine levels by 25% and vitamin B12 produced an additional 7% reduction in blood homocysteine levels, whereas vitamin B6 had no significant additive effect (30). Our findings support effects of both the folic acid and the other micronutrients in the intervention group acting to improve 1-carbon status and reduce plasma homocysteine, with potential benefit for pregnancy and offspring outcomes (31).

In the control group, plasma riboflavin and flavin mononucleotide showed modest falls from preconception to early pregnancy, and again from early to late pregnancy. We considered measuring the gold standard erythrocyte glutathione reductase activation coefficient as a specific riboflavin deficiency marker, but resource to support this could not be secured. Detailed studies have identified ≤26.5 nmol/L as the change-point of plasma riboflavin below which the erythrocyte glutathione reductase activation coefficient becomes elevated (25). Using this 26.5 nmol/L threshold, 82.0% of participants had a “marginal riboflavin status” at baseline, with 7.5% having a riboflavin concentration <5 nmol/L. In the intervention group, supplementation with 1.8 mg/day riboflavin, as in many prenatal supplements, substantially raised plasma riboflavin and reduced the prevalences of marginal and low riboflavin status before and during pregnancy. The implications of marginal or low riboflavin status before and during pregnancy have rarely been studied, but in a large cohort of pregnant women in Ireland and Northern Ireland biomarker analysis showed that 68% had low or deficient riboflavin status, which was associated with a higher risk of anaemia during pregnancy (32).

Plasma pyridoxal 5-phosphate in the control group showed a modest fall from preconception to early pregnancy, but then a substantial fall from early to late pregnancy, accompanied by a sharp rise in plasma HK and cystathionine:cysteine ratios, reflecting impairment of vitamin B6 dependent pathways in late pregnancy. For vitamin B6, pyridoxal 5-phosphate is the most commonly used status marker (33); the HK ratio, composed of HK and the four kynurenines that are products of the pyridoxal 5-phosphate dependent enzymes, kynurenine transaminase and kynureninase has been developed as a marker of tryptophan catabolism regulation by vitamin B6 that rises in B6 deficiency (33, 34). Evaluation has also found that the ratio of cystathionine:cysteine (transsulfuration pathway regulation) has merit as a B6 intake marker, while being more subject to other influences than the HK ratio (22). The marked late gestation rises in HK ratio is consistent with evidence of kynurenine pathway enzyme inhibition by the more than 10-fold physiological increase in estrogen with advancing gestation from mid to late pregnancy (35, 36). This late gestation inhibition of the kynurenine pathway may be partly physiological but nonetheless has implications for B6 nutrient supply to the fetus and potential impacts on later offspring health. Notably, there is increasing evidence linking childhood kynurenine pathway perturbations with metabolic health risk (37). In our study, the intervention, providing 2.6 mg vitamin B6 daily, substantially increased plasma pyridoxal 5-phosphate levels and lowered the plasma HK ratio in late pregnancy, with a lowering of the cystathionine:cysteine ratio also supporting a lesser impairment of vitamin B6 dependent pathways. While the US Institute of Medicine Recommended Dietary Allowance for vitamin B6 in pregnancy is 1.9 mg/day (38), European Community recommendations for pregnancy are that vitamin B6 supplements of 2.5-4 mg/day are required to maintain the plasma concentration of vitamin B6 at pre-pregnancy levels (39).

In the control group plasma vitamin B12 showed modest falls from preconception to early pregnancy, and a larger fall from early to late pregnancy, accompanied by increases in the late pregnancy prevalences of vitamin B12 deficiency and depletion (11.1% and 55.2%, respectively) and a rise in plasma methylmalonic acid as a marker of vitamin B12 insufficiency; this is in keeping with previous reports of raised vitamin B12 deficiency markers in late pregnancy (40). Supplementation with 5.2 µg/day vitamin B12 led to higher plasma cobalamin and lower methylmalonic acid during pregnancy in the intervention group. Recommended daily allowance amounts for vitamin B12 vary internationally. Meta-analysis of randomised trials in non-pregnant individuals concluded that a doubling of vitamin B12 intake increases vitamin B12 concentrations by 14%, with a consistent benefit of amounts similar to the 5.2 µg/day used in this trial (41), whereas smaller amounts are less consistent in their effect on B12 status (42). Systematic review has linked lower maternal vitamin B12 status with a variety of adverse pregnancy outcomes, including a higher risk of neural tube defects, recurrent pregnancy losses, gestational diabetes, pre-eclampsia, and lower birth weight (43), and meta-analysis has shown that lower plasma concentrations associate with a higher risk of preterm delivery (44). Lower maternal concentrations are also associated with adverse cardiometabolic and neurocognitive outcomes in the offspring (43, 45). While randomised trials of vitamin B12 supplementation in pregnancy have been inconclusive in relation to an effect on birth weight they support a beneficial effect on offspring neurocognitive development (43). In our study maternal plasma cobalamin was still higher in the intervention group post-delivery, 6-months after discontinuation of supplementation, most likely reflecting repletion of hepatic stores. It is likely this would increase breast milk vitamin B12 supply to the infant given the strong correlation between maternal plasma and breast milk concentrations (46). This is potentially of importance as post-delivery maternal vitamin B12 supplementation with the low doses found in most over-the-counter supplements (1 to 10 µg/day) are widely thought to result in only modest increases in human milk levels (47).

Plasma 25-hydroxyvitamin D in the control group taking a supplement without vitamin D was remarkably unchanged from preconception baseline to early pregnancy, then increased modestly between early and late pregnancy, with high prevalences of vitamin D deficiency and insufficiency at baseline preconception (47.7% and 85.7%, respectively) and through pregnancy. Supplementation with 10 µg of vitamin D in the intervention group led to progressive increases in plasma 25-hydroxyvitamin D and a substantial reduction in the prevalences of vitamin D deficiency and insufficiency during pregnancy (e.g. 8.5% and 27.3% in late pregnancy, respectively vs 35.1% and 64.6% in the control group). The dose of vitamin D recommended for routine use in pregnancy varies internationally, with some advocating 100 µg daily or higher (48), while the World Health Organization/Food and Agriculture Organization of the United Nations recommends 5 µg daily (49); in the UK it is recommended that pregnant women should consider taking a supplement containing 10 µg between September and March (4). In the MAVIDOS randomised trial supplementation 25 µg of vitamin D daily from 14 weeks gestation until delivery lowered the incidence of infantile atopic eczema (5) and increased childhood areal bone mineral density in the offspring (6). Vitamin D is stored in adipose tissue and our trial suggests that low dose (10 µg) supplementation over a long period starting preconception can support improved gestational vitamin D status.

The finding of significant prevalences of vitamin insufficiencies in women living in high income countries who are attempting to become pregnant is a serious concern. This is particularly the case given increasing advocacy to reduce meat and dairy intakes to achieve “net-zero” carbon emissions, as these foods are micronutrient dense and support nutritional adequacy (50, 51). The high prevalence of vitamin insufficiencies and increasing move towards plant-based diets, which lack vitamin B12 and are low in other micronutrients, is likely to result in more women resorting to over-the-counter supplements. Currently, over-the-counter supplements intended for preconception and pregnancy show considerable variation in which micronutrients they include and in the amounts of those included. As the amounts incorporated into the intervention supplement were comparable to those available in over-the-counter supplements, the effects we report on vitamin status before and during pregnancy are generalizable and fill an important gap in the literature.

To date changes in vitamin status from preconception to early and late pregnancy and postpartum have largely been inferred from cross-sectional data, with lower concentrations during pregnancy often ascribed to plasma volume expansion (12, 13). Our findings show however that the magnitude and pattern of change varies between nutrients, inconsistent with an effect wholly due to physiological hemodilution, and that markers of functional B6 (HK ratio) and B12 (methylmalonic acid) insufficiency increase during pregnancy. Variations between individual vitamins in maternal metabolism, intake, renal loss and feto-placental demands during pregnancy may all contribute to these varying patterns. Our randomized trial has shown for the first time that preconception and pregnancy supplementation including riboflavin, folic acid and vitamins B6, B12 and D in amounts available in over-the-counter supplements can contribute to the reduction in vitamin insufficiencies during the preconception, pregnancy and lactational periods. The potential benefits for pregnancy outcome and offspring health remain to be characterized.

### Strengths and Limitations

Strengths of our study include longitudinal plasma samples from preconception through pregnancy to 6-months post-delivery, an interventional component, a relatively large sample size and inclusion of multiple ethnic groups. The robust conduct of this double-blind RCT, which included prospectively collected data and minimization of residual confounding through randomization, with external oversight by an independent data monitoring and trial steering committee, is a strength. Over 96% of women showed good adherence defined *a priori* as supplement intake greater than 60% averaged from recruitment to delivery. Even though recruitment occurred across three different countries with inclusion of multiple ethnicities, generalizability to the global population is limited by the lack of African and Amerindian women in particular. The recruited population were generally healthy and well-nourished, yet evidence recognized as indicating micronutrient insufficiency was widespread; whether some changes represent a normal physiological change in pregnancy or a true insufficiency awaits characterization of relations with pregnancy and offspring outcomes. Another strength is the use of an accredited clinical laboratory for analyses of clinical biomarkers. In this study the entire sample set was analyzed for tryptophan and kynurenine metabolites in a single laboratory by a targeted LC-MS/MS method including authentic labelled internal standards for each analyte providing high analytical precision. The analyses were carried out in continuous sample batches, with small variability between each batch. Finally, standardized scores were used in the regression models so that the strength of associations were comparable.

### Conclusions

Significant proportions of preconception women living in high income countries have marginal or low status of folate, riboflavin, vitamin B12 and vitamin D, and many develop markers of vitamin B6 deficiency in late pregnancy. In the absence of the intervention supplement, maternal plasma concentrations show differing longitudinal patterns between vitamins from preconception to early and late pregnancy, inconsistent with plasma volume expansion wholly accounting for lower gestational concentrations, and markers of functional B6 and B12 insufficiency increase during pregnancy. Preconception and pregnancy supplementation in amounts available in over-the-counter supplements substantially reduced the prevalence of deficiency/depletion markers before and during pregnancy, and a higher maternal plasma vitamin B12 was maintained during the recommended lactational period. In the setting of increasing advocacy for more diets that are likely to be less nutrient dense, the findings suggest a need to reappraise dietary recommendations for preconception and pregnancy and to consider further the role of multiple micronutrient supplements in women living in higher income countries.

## Data Availability

Individual participant data may be shared upon reasonable requests and are subject to approval by the trial management group and the trial consultative panel. Application can be made through the corresponding author.

## Author Contributions

**Keith M Godfrey:** Conceptualization, Methodology, Investigation, Writing review and editing, Supervision, Project administration, Funding acquisition. **Philip Titcombe:** Validation, Analysis, Data curation, Writing original draft, Visualization; **Sarah El-Heis:** Methodology, Investigation, Writing review and editing; **Benjamin Albert:** Investigation, Writing review and editing; **Elizabeth Tham:** Investigation, Writing review and editing; **Sheila Barton:** Methodology, Validation, Analysis, Resources, Data curation, Writing review and editing; **Timothy Kenealy:** Investigation, Writing review and editing; **Heidi Nield:** Resources, Project administration, Writing review and editing; **Mary Foong-Fong Chong:** Writing review and editing; **Yap Seng Chong:** Conceptualization, Methodology, Writing review and editing, Supervision, Funding acquisition; **Shiao-Yng Chan:** Conceptualization, Methodology, Validation, Analysis, Investigation, Writing original draft, Visualization, Supervision, Project administration; **Wayne Cutfield:** Methodology, Investigation, Writing review and editing, Supervision, Project administration;

## Acknowledgements

The NiPPeR Study Group authors for the Medline citation comprises the following:

*National University Hospital, Singapore:* Julie Ann Guiao Castro, Wendy Sim, Gladys Woon, Hsin Fang Chang

*Singapore Institute for Clinical Sciences, Agency for Science, Technology and Research, Singapore*: Gernalia Satianegara, Karen ML Tan, Vicky Tay, Jui-Tsung Wong

*MRC Lifecourse Epidemiology Centre, University of Southampton, UK*: Paula Costello, Vanessa Cox, Nicholas C Harvey, Sevasti Galani

*Liggins Institute, University of Auckland, New Zealand*: Mary Cavanagh, Judith Hammond, Mark H. Vickers

*Société Des Produits Nestlé S.A.*: Aristea Binea, Irma Silva-Zolezzi, Chiara Nembrini

We thank the participants and their families for their enthusiastic involvement in the study; the study research staff and hospital clinical staff at participating centers, and operational support staff for their contributions to the trial; and the members of the Independent Data Monitoring and Safety Committee for their contributions and oversight of the conduct of the trial.

## Abbreviations list

AA: anthranilic acid
BMI: body mass index
CI: confidence interval
HAA: hydroxyanthranilic acid
HIV: human immunodeficiency virus
HK: 3-hydroxykynurenine
KA: kynurenic acid
LC-MS/MS: Liquid chromatography with tandem mass spectrometry
NiPPeR Trial: Nutritional Intervention Preconception and During Pregnancy to Maintain Healthy Glucose Metabolism and Offspring Health trial
NRES: National Research Ethics Service
SDS: standard deviation score
UK: United Kingdom
US: United States
XA: 3-xanthurenic acid

## Clinical trial registration

1) Date of registration: 15^th^ July 2015
2) Date of initial participant enrollment: 3^rd^ August 2015
3) Clinical trial identification number: NCT02509988; U1111-1171-8056
4) URL of the registration site: https://www.clinicaltrials.gov/ct2/show/NCT02509988
5) Data sharing information: Individual participant data may be shared upon reasonable requests and are subject to approval by the trial management group and the trial consultative panel. Application can be made through the corresponding author.

---

**Supplementary Figure 1.**
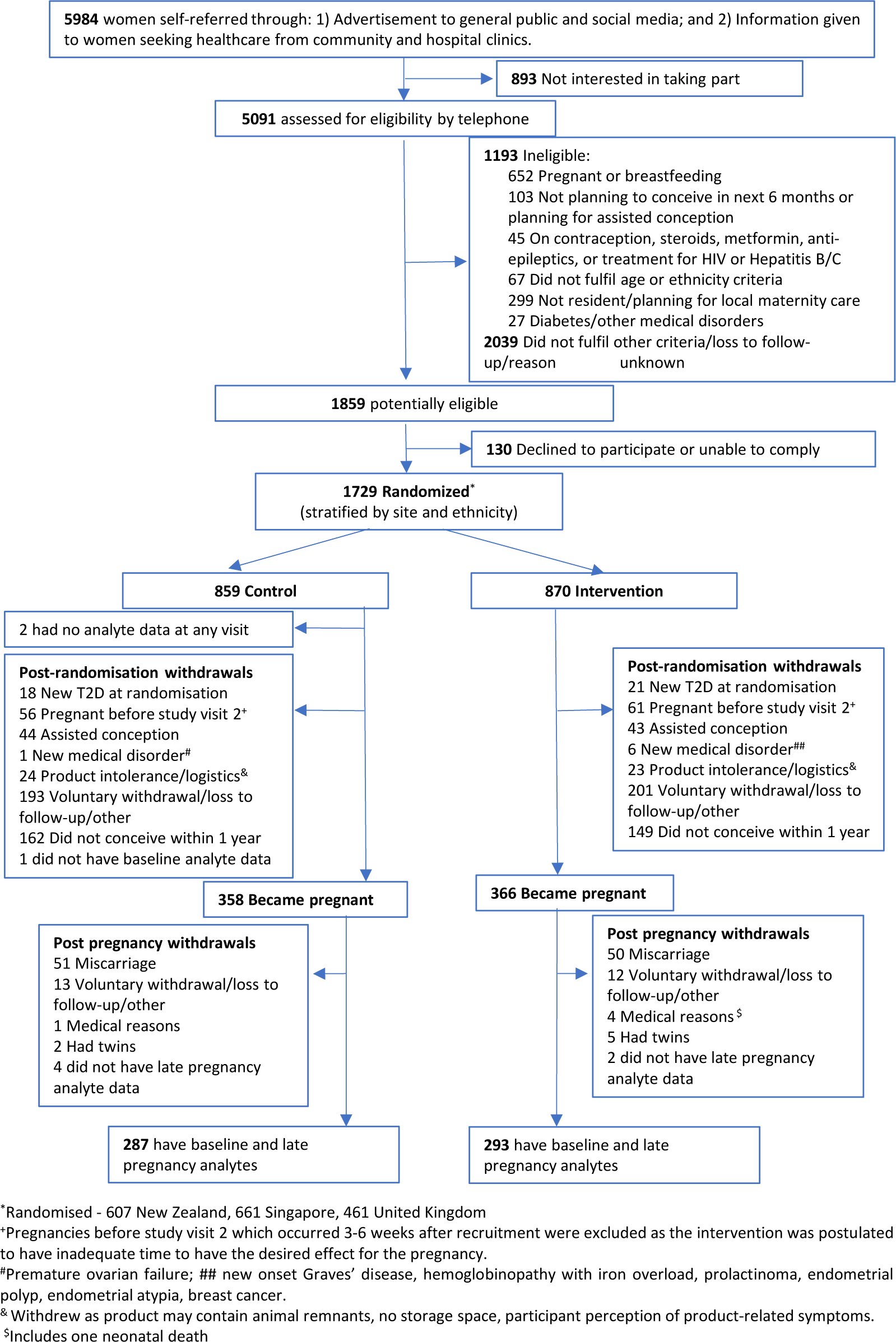
Flowchart of study participants from assessment of eligibility, through randomization, conception, pregnancy and post-delivery. Abbreviation: Cesarean, cesarean section delivery.

**Supplementary Figure 2.**
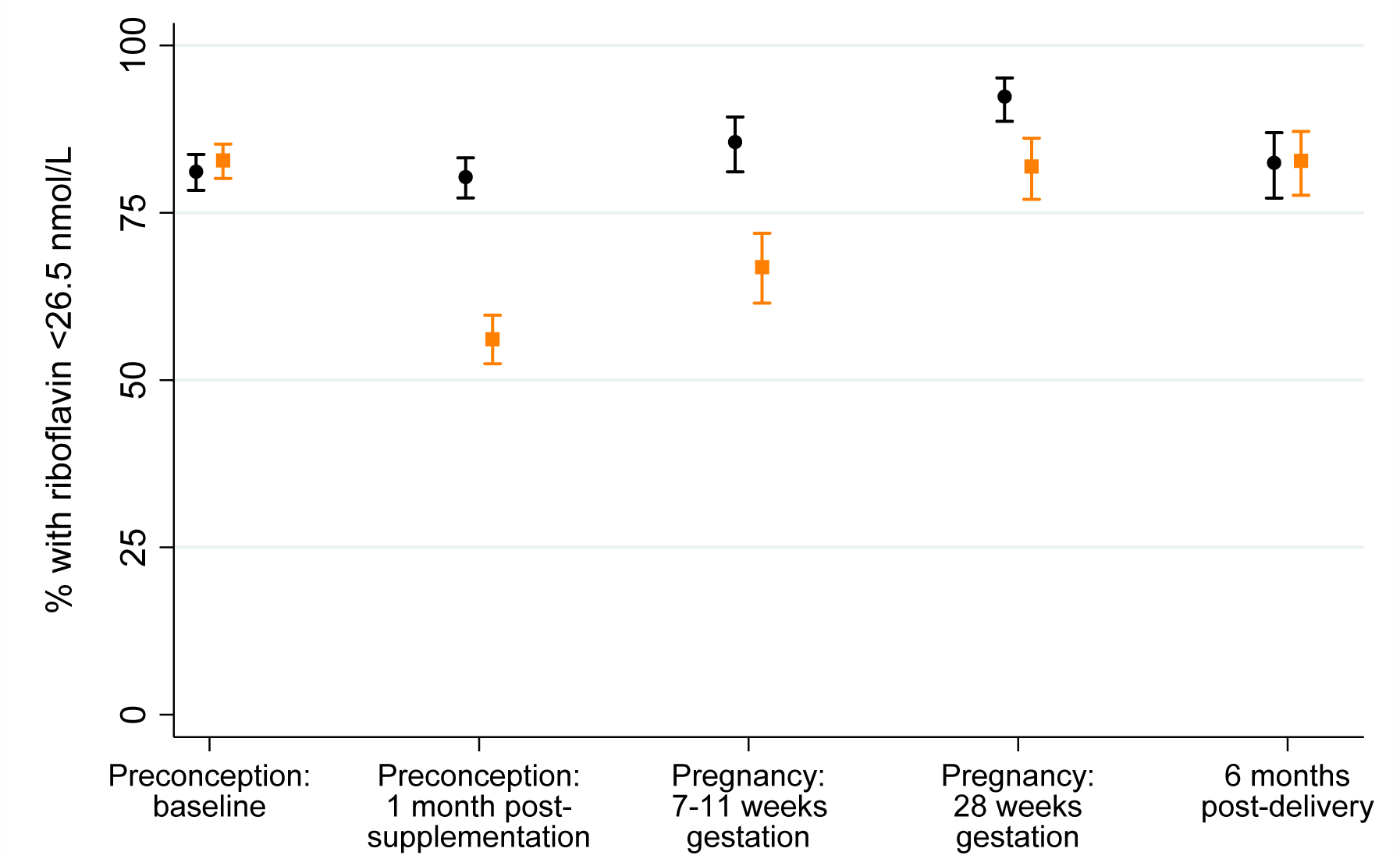
Marginal riboflavin status (plasma riboflavin ≤26.5 nmol/L)

**Supplementary Figure 3.**
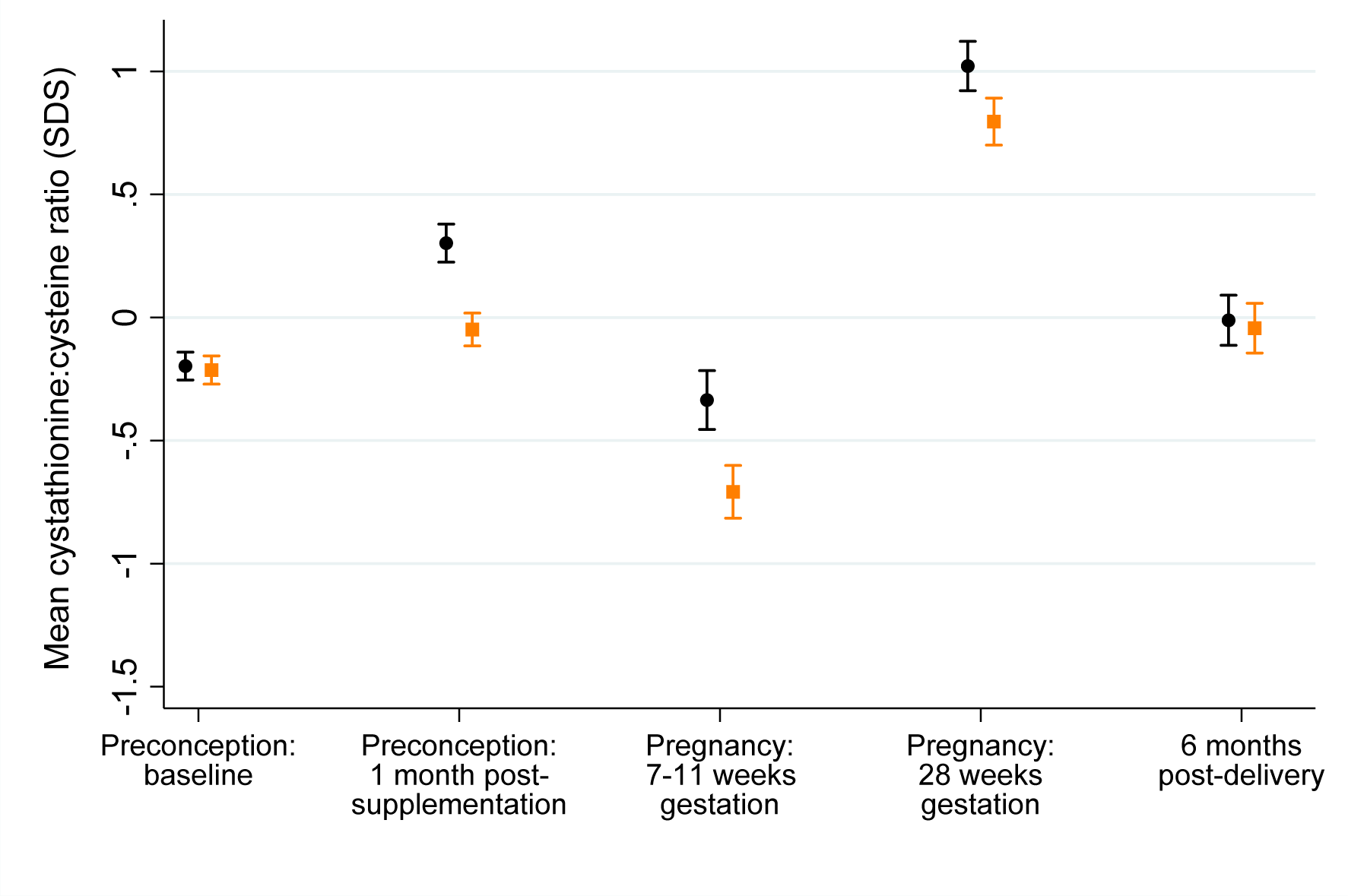
Plasma cystathionine/cysteine ratio.

**Supplementary Figure 4.**
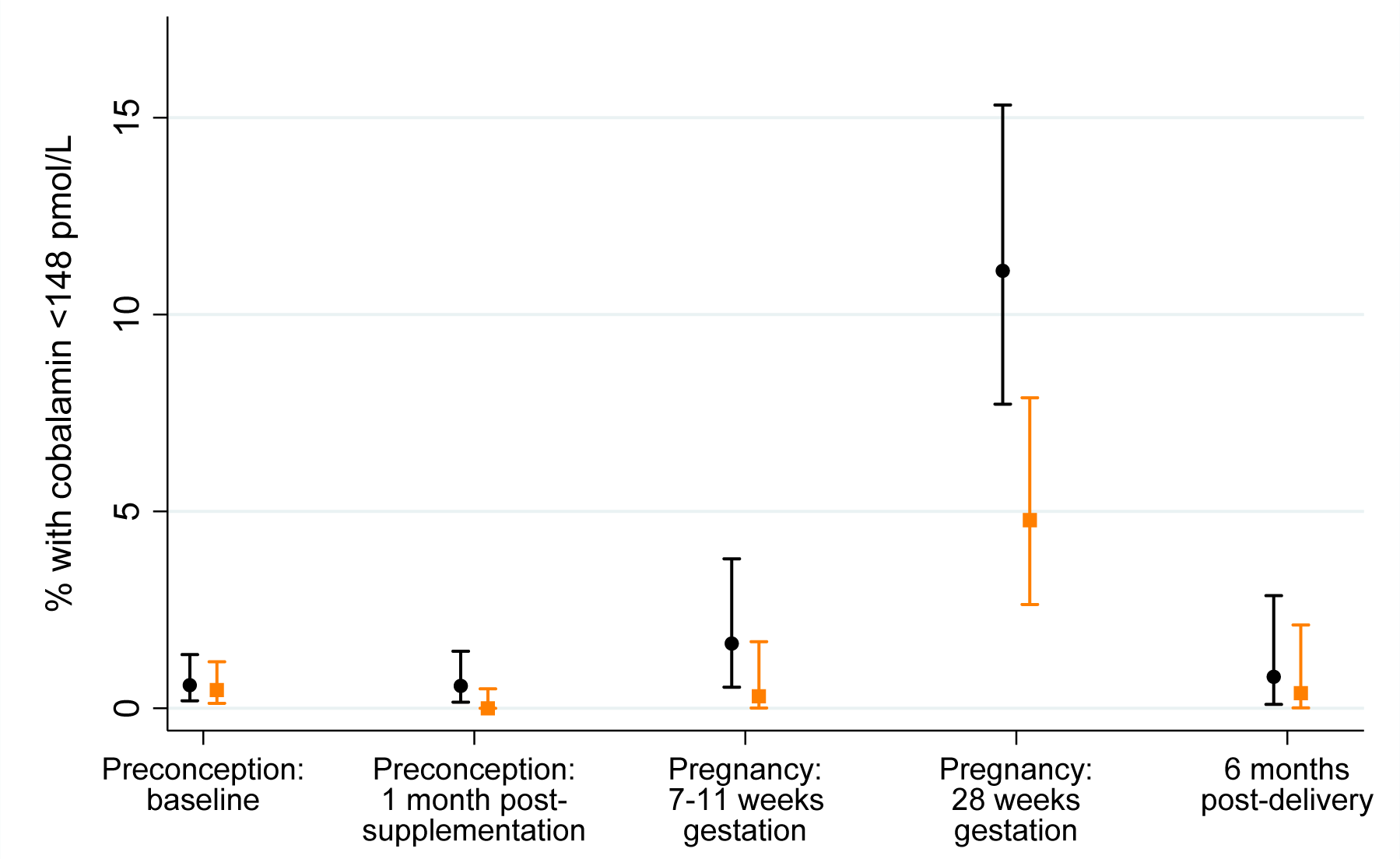
Vitamin B12 deficiency (<148 pmol/L)

**Supplementary Table 1:** Median (IQR) values in original units according to control or intervention group at each time-point, for participants with measurements at each of preconception baseline and late pregnancy.

|  | Preconception baseline |  | Preconception 1-month post supplementation |  | Early pregnancy (7-11 weeks gestation) |  | Late pregnancy (28 weeks gestation) |  | 6 months post-delivery |  |
| --- | --- | --- | --- | --- | --- | --- | --- | --- | --- | --- |
|  | Controls (n=287) | Intervention (n=293) | Controls (n=286) | Intervention (n=292) | Controls (n=276) | Intervention (n=289) | Controls (n=287) | Intervention (n=293) | Controls (n=247) | Intervention (n=260) |
| Folate (nmol/L) | 23.1<br>(13.5, 41.3) | 23.8<br>(13.6, 42.6) | 44.4<br>(30.9, 56.3) | 42.9<br>(31.4, 54.2) | 49.9<br>(40.6, 60.2) | 52.2<br>(43.7, 63.0) | 41.5<br>(30.6, 51.4) | 44.1<br>(34.3, 54.5) | 19.3<br>(11.9, 33.2) | 19.0<br>(12.4, 32.4) |
| Homocysteine (μmol/L) | 7.0<br>(6.1, 8.3) | 7.1<br>(6.0, 8.4) | 6.8<br>(6.0, 7.9) | 6.3<br>(5.5, 7.3) | 5.5<br>(4.8, 6.4) | 4.8<br>(4.2, 5.5) | 4.6<br>(3.9, 5.2) | 3.9<br>(3.5, 4.6) | 7.4<br>(6.4, 9.0) | 7.2<br>(6.1, 8.5) |
| Riboflavin (nmol/L) | 12.7<br>(7.5, 19.6) | 13.7<br>(8.6, 21.3) | 12.6<br>(8.4, 21.5) | 25.1<br>(16.8, 38.0) | 10.8<br>(6.7, 18.4) | 19.7<br>(13.0, 29.8) | 10.2<br>(6.6, 15.1) | 16.9<br>(12.2, 23.3) | 13.0<br>(8.9, 21.1) | 12.8<br>(8.4, 22.6) |
| FMN (nmol/L) | 14.7<br>(11.4, 18.7) | 15.0<br>(12.0, 19.6) | 15.6<br>(12.7, 19.5) | 19.0<br>(15.1, 25.1) | 14.3<br>(11.4, 18.3) | 17.4<br>(14.2, 21.6) | 10.1<br>(8.4, 12.0) | 11.5<br>(9.8, 13.4) | 14.2<br>(11.8, 17.0) | 14.4<br>(11.2, 18.7) |
| Pyridoxal 5-phosphate (nmol/L) | 59.4<br>(42.4, 92.1) | 61.0<br>(44.9, 99.0) | 57.2<br>(44.7, 79.3) | 144<br>(107, 187) | 47.6<br>(34.7, 64.2) | 104.0<br>(75.8, 138.0) | 19.2<br>(14.6, 28.3) | 41.9<br>(30.3, 56.4) | 55.9<br>(40.0, 88.1) | 56.4<br>(40.5, 86.8) |
| HK ratio (no units) | 0.35<br>(0.30, 0.43) | 0.36<br>(0.30, 0.41) | 0.36<br>(0.29, 0.42) | 0.29<br>(0.26, 0.34) | 0.35<br>(0.28, 0.42) | 0.29<br>(0.24, 0.33) | 0.51<br>(0.41,0.61) | 0.44<br>(0.37,0.54) | 0.40<br>(0.32, 0.49) | 0.39<br>(0.32, 0.47) |
| Cobalamin (pmol/L) | 355<br>(276, 444) | 354<br>(278, 434) | 355<br>(278, 449) | 447<br>(357, 553) | 307<br>(240, 385) | 411<br>(332, 511) | 214<br>(176, 270) | 299<br>(248, 380) | 342<br>(265, 438) | 386<br>(312, 466) |
| MMA (μmol/L) | 0.13<br>(0.11, 0.17) | 0.14<br>(0.11, 0.18) | 0.15<br>(0.12, 0.21) | 0.15<br>(0.12, 0.19) | 0.12<br>(0.10, 0.15) | 0.11<br>(0.10, 0.14) | 0.14<br>(0.11,0.20) | 0.12<br>(0.10,0.15) | 0.15<br>(0.12, 0.20) | 0.14<br>(0.12, 0.17) |
| Vitamin D3 (nmol/L) | 53.5<br>(40.3, 68.0) | 54.4<br>(39.6, 70.0) | 54.4<br>(40.5, 69.4) | 64.8<br>(54.5, 76.4) | 52.8<br>(41.5, 69.4) | 70.0<br>(57.6, 81.8) | 62.9<br>(41.0, 87.5) | 92.8<br>(73.0,106.9) | 62.3<br>(45.3, 77.6) | 65.1<br>(48.4, 78.2) |

HK ratio – 3'-hydroxykynurenine ratio, FMN – flavin mononucleotide, MMA - methylmalonic acid

## References

1. Keats EC, Haider BA, Tam E, Bhutta ZA. Multiple-micronutrient supplementation for women during pregnancy. The Cochrane database of systematic reviews. 2019;3(3):Cd004905.

2. Prevention of neural tube defects: results of the Medical Research Council Vitamin Study. MRC Vitamin Study Research Group. Lancet (London, England). 1991;338(8760):131–7.

3. Cooper C, Harvey NC, Bishop NJ, Kennedy S, Papageorghiou AT, Schoenmakers I, et al. Maternal gestational vitamin D supplementation and offspring bone health (MAVIDOS): a multicentre, double-blind, randomised placebo-controlled trial. The lancet Diabetes & endocrinology. 2016;4(5):393–402.

4. NHS. [Available from: https://www.nhs.uk/pregnancy/keeping-well/vitamins-supplements-and-nutrition/.

5. El-Heis S, D’Angelo S, Curtis EM, Healy E, Moon RJ, Crozier SR, et al. Maternal antenatal vitamin D supplementation and offspring risk of atopic eczema in the first 4 years of life: evidence from a randomized controlled trial. The British journal of dermatology. 2022;187(5):659–66.

6. Curtis EM, Moon RJ, D’Angelo S, Crozier SR, Bishop NJ, Gopal-Kothandapani JS, et al. Pregnancy Vitamin D Supplementation and Childhood Bone Mass at Age 4 Years: Findings From the Maternal Vitamin D Osteoporosis Study (MAVIDOS) Randomized Controlled Trial. JBMR plus. 2022;6(7):e10651.

7. Fleming TP, Watkins AJ, Velazquez MA, Mathers JC, Prentice AM, Stephenson J, et al. Origins of lifetime health around the time of conception: causes and consequences. Lancet (London, England). 2018;391(10132):1842–52.

8. Stephenson J, Heslehurst N, Hall J, Schoenaker D, Hutchinson J, Cade JE, et al. Before the beginning: nutrition and lifestyle in the preconception period and its importance for future health. Lancet (London, England). 2018;391(10132):1830–41.

9. Ronnenberg AG, Goldman MB, Chen D, Aitken IW, Willett WC, Selhub J, et al. Preconception homocysteine and B vitamin status and birth outcomes in Chinese women. Am J Clin Nutr. 2002;76(6):1385–91.

10. Robinson SM, Crozier SR, Miles EA, Gale CR, Calder PC, Cooper C, et al. Preconception Maternal Iodine Status Is Positively Associated with IQ but Not with Measures of Executive Function in Childhood. The Journal of nutrition. 2018;148(6):959–66.

11. Sukumar N, Adaikalakoteswari A, Venkataraman H, Maheswaran H, Saravanan P. Vitamin B_12_ status in women of childbearing age in the UK and its relationship with national nutrient intake guidelines: results from two National Diet and Nutrition Surveys. BMJ Open. 2016;6(8):e011247.

12. Aguree S, Gernand AD. Plasma volume expansion across healthy pregnancy: a systematic review and meta-analysis of longitudinal studies. BMC pregnancy and childbirth. 2019;19(1):508.

13. Hall MH, Pirani BB, Campbell D. The cause of the fall in serum folate in normal pregnancy. British journal of obstetrics and gynaecology. 1976;83(2):132–6.

14. Sahariah SA, Gandhi M, Chopra H, Kehoe SH, Johnson MJ, di Gravio C, et al. Body Composition and Cardiometabolic Risk Markers in Children of Women who Took Part in a Randomized Controlled Trial of a Preconceptional Nutritional Intervention in Mumbai, India. The Journal of nutrition. 2022;152(4):1070–81.

15. D’Souza N, Behere RV, Patni B, Deshpande M, Bhat D, Bhalerao A, et al. Pre-conceptional Maternal Vitamin B12 Supplementation Improves Offspring Neurodevelopment at 2 Years of Age: PRIYA Trial. Frontiers in pediatrics. 2021;9:755977.

16. Godfrey KM, Cutfield W, Chan SY, Baker PN, Chong YS, NiPPeR Study Group. Nutritional Intervention Preconception and During Pregnancy to Maintain Healthy Glucose Metabolism and Offspring Health (“NiPPeR”): study protocol for a randomised controlled trial. Trials. 2017;18(1):131.

17. Godfrey KM, Barton SJ, El-Heis S, Kenealy T, Nield H, Baker PN, et al. Myo-Inositol, Probiotics, and Micronutrient Supplementation From Preconception for Glycemia in Pregnancy: NiPPeR International Multicenter Double-Blind Randomized Controlled Trial. Diabetes care. 2021;44(5):1091–9.

18. Zhang H, Lv Y, Li Z, Sun L, Guo W. The efficacy of myo-inositol supplementation to prevent gestational diabetes onset: a meta-analysis of randomized controlled trials. The journal of maternal-fetal & neonatal medicine : the official journal of the European Association of Perinatal Medicine, the Federation of Asia and Oceania Perinatal Societies, the International Society of Perinatal Obstet. 2019;32(13):2249–55.

19. Laitinen K, Poussa T, Isolauri E. Probiotics and dietary counselling contribute to glucose regulation during and after pregnancy: a randomised controlled trial. The British journal of nutrition. 2009;101(11):1679–87.

20. Midttun Ø, Hustad S, Ueland PM. Quantitative profiling of biomarkers related to B-vitamin status, tryptophan metabolism and inflammation in human plasma by liquid chromatography/tandem mass spectrometry. Rapid communications in mass spectrometry : RCM. 2009;23(9):1371–9.

21. Ulvik A, McCann A, Midttun Ø, Meyer K, Godfrey KM, Ueland PM. Quantifying Precision Loss in Targeted Metabolomics Based on Mass Spectrometry and Nonmatching Internal Standards. Analytical chemistry. 2021;93(21):7616–24.

22. Clasen JL, Heath AK, Van Puyvelde H, Huybrechts I, Park JY, Ferrari P, et al. A comparison of complementary measures of vitamin B6 status, function, and metabolism in the European Prospective Investigation into Cancer and Nutrition (EPIC) study. Am J Clin Nutr. 2021;114(1):338–47.

23. Oner N, Vatansever U, Karasalihoğlu S, Ekuklu G, Celtik C, Biner B. The prevalence of folic acid deficiency among adolescent girls living in Edirne, Turkey. The Journal of adolescent health : official publication of the Society for Adolescent Medicine. 2006;38(5):599–606.

24. Moretti R, Caruso P. The Controversial Role of Homocysteine in Neurology: From Labs to Clinical Practice. International journal of molecular sciences. 2019;20(1).

25. Tan A, Zubair M, Ho C-l, McAnena L, McNulty H, Ward M, et al. Plasma riboflavin concentration as novel indicator for vitamin-B2 status assessment: suggested cutoffs and its association with vitamin-B6 status in women. Proceedings of the Nutrition Society. 2020;79(OCE2):E658.

26. Simpson JL, Bailey LB, Pietrzik K, Shane B, Holzgreve W. Micronutrients and women of reproductive potential: required dietary intake and consequences of dietary deficiency or excess. Part I--Folate, Vitamin B12, Vitamin B6. The journal of maternal-fetal & neonatal medicine : the official journal of the European Association of Perinatal Medicine, the Federation of Asia and Oceania Perinatal Societies, the International Society of Perinatal Obstet. 2010;23(12):1323–43.

27. Loo EXL, Tham EH, Phang KW, Goh A, Teoh OH, Chong YS, et al. Associations between maternal vitamin D levels during pregnancy and allergic outcomes in the offspring in the first 5 years of life. Pediatric allergy and immunology : official publication of the European Society of Pediatric Allergy and Immunology. 2019;30(1):117–22.

28. Ducker GS, Rabinowitz JD. One-Carbon Metabolism in Health and Disease. Cell metabolism. 2017;25(1):27–42.

29. Ducros V, Andriollo-Sanchez M, Arnaud J, Meunier N, Laporte F, Hininger-Favier I, et al. Zinc supplementation does not alter plasma homocysteine, vitamin B12 and red blood cell folate concentrations in French elderly subjects. Journal of trace elements in medicine and biology : organ of the Society for Minerals and Trace Elements (GMS). 2009;23(1):15–20.

30. Clarke R, Armitage J. Vitamin supplements and cardiovascular risk: review of the randomized trials of homocysteine-lowering vitamin supplements. Seminars in thrombosis and hemostasis. 2000;26(3):341–8.

31. Rojas-Gómez A, Solé-Navais P, Cavallé-Busquets P, Ornosa-Martin G, Grifoll C, Ramos-Rodriguez C, et al. Pregnancy homocysteine and cobalamin status predict childhood metabolic health in the offspring. Pediatric research. 2022.

32. Duffy B, Pentieva K, Ward M, Psara E, O’Sullivan E, Horrigan G, et al. Impact of riboflavin status on haemoglobin and risk of anaemia in pregnancy. Proceedings of the Nutrition Society. 2021;80(OCE1):E45.

33. Ueland PM, Ulvik A, Rios-Avila L, Midttun Ø, Gregory JF. Direct and Functional Biomarkers of Vitamin B6 Status. Annual review of nutrition. 2015;35:33–70.

34. Ulvik A, Midttun Ø, McCann A, Meyer K, Tell G, Nygård O, et al. Tryptophan catabolites as metabolic markers of vitamin B-6 status evaluated in cohorts of healthy adults and cardiovascular patients. Am J Clin Nutr. 2020;111(1):178–86.

35. Jayawickrama GS, Nematollahi A, Sun G, Gorrell MD, Church WB. Inhibition of human kynurenine aminotransferase isozymes by estrogen and its derivatives. Sci Rep. 2017;7(1):17559.

36. Abbassi-Ghanavati M, Greer LG, Cunningham FG. Pregnancy and laboratory studies: a reference table for clinicians. Obstetrics and gynecology. 2009;114(6):1326–31.

37. Tan KM, Tint MT, Kothandaraman N, Yap F, Godfrey KM, Lee YS, et al. Association of plasma kynurenine pathway metabolite concentrations with metabolic health risk in prepubertal Asian children. International journal of obesity (2005). 2022;46(6):1128–37.

38. Institute of Medicine Standing Committee on the Scientific Evaluation of Dietary Reference I, its Panel on Folate OBV, Choline. The National Academies Collection: Reports funded by National Institutes of Health. Dietary Reference Intakes for Thiamin, Riboflavin, Niacin, Vitamin B(6), Folate, Vitamin B(12), Pantothenic Acid, Biotin, and Choline. Washington (DC): National Academies Press (US) Copyright © 1998, National Academy of Sciences.; 1998.

39. Proposed nutrient and energy intakes for the European community: a report of the Scientific Committee for Food of the European community. Nutrition reviews. 1993;51(7):209-12.

40. Greibe E, Andreasen BH, Lildballe DL, Morkbak AL, Hvas AM, Nexo E. Uptake of cobalamin and markers of cobalamin status: a longitudinal study of healthy pregnant women. Clinical chemistry and laboratory medicine. 2011;49(11):1877–82.

41. Dullemeijer C, Souverein OW, Doets EL, van der Voet H, van Wijngaarden JP, de Boer WJ, et al. Systematic review with dose-response meta-analyses between vitamin B-12 intake and European Micronutrient Recommendations Aligned’s prioritized biomarkers of vitamin B-12 including randomized controlled trials and observational studies in adults and elderly persons. Am J Clin Nutr. 2013;97(2):390–402.

42. Sanchez H, Albala C, Lera L, Dangour AD, Uauy R. Effectiveness of the National Program of Complementary Feeding for older adults in Chile on vitamin B12 status in older adults; secondary outcome analysis from the CENEX Study (ISRCTN48153354). Nutrition journal. 2013;12:124.

43. Behere RV, Deshmukh AS, Otiv S, Gupte MD, Yajnik CS. Maternal Vitamin B12 Status During Pregnancy and Its Association With Outcomes of Pregnancy and Health of the Offspring: A Systematic Review and Implications for Policy in India. Frontiers in endocrinology. 2021;12:619176.

44. Rogne T, Tielemans MJ, Chong MF, Yajnik CS, Krishnaveni GV, Poston L, et al. Associations of Maternal Vitamin B12 Concentration in Pregnancy With the Risks of Preterm Birth and Low Birth Weight: A Systematic Review and Meta-Analysis of Individual Participant Data. American journal of epidemiology. 2017;185(3):212–23.

45. Lai JS, Mohamad Ayob MN, Cai S, Quah PL, Gluckman PD, Shek LP, et al. Maternal plasma vitamin B12 concentrations during pregnancy and infant cognitive outcomes at 2 years of age. The British journal of nutrition. 2019;121(11):1303–12.

46. Bae S, West AA, Yan J, Jiang X, Perry CA, Malysheva O, et al. Vitamin B-12 Status Differs among Pregnant, Lactating, and Control Women with Equivalent Nutrient Intakes. The Journal of nutrition. 2015;145(7):1507–14.

47. Allen LH, Hampel D, Shahab-Ferdows S, York ER, Adair LS, Flax VL, et al. Antiretroviral therapy provided to HIV-infected Malawian women in a randomized trial diminishes the positive effects of lipid-based nutrient supplements on breast-milk B vitamins. Am J Clin Nutr. 2015;102(6):1468–74.

48. Pérez-López FR, Pilz S, Chedraui P. Vitamin D supplementation during pregnancy: an overview. Current Opinion in Obstetrics and Gynecology. 2020;32(5):316–21.

49. Organization WH. Food and agriculture organization of the United Nations. Vitamin and mineral requirements in human nutrition. 2004;2:17–299.

50. Neufingerl N, Eilander A. Nutrient Intake and Status in Adults Consuming Plant-Based Diets Compared to Meat-Eaters: A Systematic Review. Nutrients. 2021;14(1).

51. Hobbs DA, Durrant C, Elliott J, Givens DI, Lovegrove JA. Diets containing the highest levels of dairy products are associated with greater eutrophication potential but higher nutrient intakes and lower financial cost in the United Kingdom. European journal of nutrition. 2020;59(3):895–908.

